# An Evaluation of Prospective COVID-19 Modeling: From Data to Science Translation

**DOI:** 10.1101/2022.04.18.22273992

**Authors:** Kristen Nixon, Sonia Jindal, Felix Parker, Nicholas G. Reich, Kimia Ghobadi, Elizabeth C. Lee, Shaun Truelove, Lauren Gardner

## Abstract

**Background:** Infectious disease modeling can serve as a powerful tool for science-based management of outbreaks, providing situational awareness and decision support for policy makers. Predictive modeling of an emerging disease is challenging due to limited knowledge on its epidemiological characteristics. For COVID-19, the prediction difficulty was further compounded by continuously changing policies, varying behavioral responses, poor availability and quality of crucial datasets, and the variable influence of different factors as the pandemic progresses. Due to these challenges, predictive modeling for COVID-19 has earned a mixed track record.

**Methods:** We provide a systematic review of prospective, data-driven modeling studies on population-level dynamics of COVID-19 in the US and conduct a quantitative assessment on crucial elements of modeling, with a focus on the aspects of modeling that are critical to make them useful for decision-makers. For each study, we documented the forecasting window, methodology, prediction target, datasets used, geographic resolution, whether they expressed quantitative uncertainty, the type of performance evaluation, and stated limitations. We present statistics for each category and discuss their distribution across the set of studies considered. We also address differences in these model features based on fields of study.

**Findings:** Our initial search yielded 2,420 papers, of which 119 published papers and 17 preprints were included after screening. The most common datasets relied upon for COVID-19 modeling were counts of cases (93%) and deaths (62%), followed by mobility (26%), demographics (25%), hospitalizations (12%), and policy (12%). Our set of papers contained a roughly equal number of short-term (46%) and long-term (60%) predictions (defined as a prediction horizon longer than 4 weeks) and statistical (43%) versus compartmental (47%) methodologies. The target variables used were predominantly cases (89%), deaths (52%), hospitalizations (10%), and *R*_*t*_ (9%). We found that half of the papers in our analysis did not express quantitative uncertainty (50%). Among short-term prediction models, which can be fairly evaluated against truth data, 25% did not conduct any performance evaluation, and most papers were not evaluated over a timespan that includes varying epidemiological dynamics. The main categories of limitations stated by authors were disregarded factors (39%), data quality (28%), unknowable factors (26%), limitations specific to the methods used (22%), data availability (16%), and limited generalizability (8%). 36% of papers did not list any limitations in their discussion or conclusion section.

**Interpretation:** Published COVID-19 models were found to be consistently lacking in some of the most important elements required for usability and translation, namely transparency, expressing uncertainty, performance evaluation, stating limitations, and communicating appropriate interpretations. Adopting the EPIFORGE 2020 guidelines would address these shortcomings and improve the consistency, reproducibility, comparability, and quality of epidemic forecasting reporting. We also discovered that most of the operational models that have been used in real-time to inform decision-making have not yet made it into the published literature, which highlights that the current publication system is not suited to the rapid information-sharing needs of outbreaks. Furthermore, data quality was identified to be one of the most important drivers of model performance, and a consistent limitation noted by the modeling community. The US public health infrastructure was not equipped to provide timely, high-quality COVID-19 data, which is required for effective modeling. Thus, a systematic infrastructure for improved data collection and sharing should be a major area of investment to support future pandemic preparedness.

## Introduction

The COVID-19 pandemic has become an unprecedented public health crisis in its prolonged impact on health and its disruption to economic and social life, with more than 5 million deaths globally as of November 2021. To aid planning and response efforts during a pandemic, mathematical modeling of current and future trends of outbreaks has historically served as a valuable tool. Nowcasting and forecasting models can improve situational awareness of the current and near future states of disease spread, while long-term projections and scenario modeling can shed light on outcomes that may result from a set of assumptions. Insights from modeling can educate individuals on how to mitigate their own risks, while also providing decision support for policy makers seeking to minimize harm to an entire population. However, despite the influx of expertise from many fields, the modeling community has put up a mixed track-record on accurately capturing COVID-19 risk in real-time.

Efforts to date have revealed some of the challenges with predicting the course of the COVID-19 pandemic^1,2^. Critically, individual risk reduction behaviors and policy compliance, which directly impact case growth, are not easily measured. New variants emerge that change the course of the pandemic. Factors such as these, coupled with a substantial lack of quality and timely data, makes it difficult to build a model that predicts accurately into the future. For some endemic infectious diseases such as influenza, modelers have shown moderate success in making short-term predictions^3,4^. Influenza predictions in the US benefit from more than 15 years of training data, more predictable new strains, and an absence of large-scale behavioral changes and policy interventions. However, even anomalous influenza seasons, like 2017/2018 in the US, have proven hard to predict. While modeling has been used for many previous emerging and endemic infectious diseases, the COVID-19 pandemic thrust modeling into the spotlight. As a result of these challenges, the utility of COVID-19 models for informing response efforts has been criticized^5–7^, largely due to a few particularly erroneous projections at the start of the outbreak.

Due to the rapid pace of publication on preprint servers and in publications, the several COVID-19 modeling reviews that have been published to date form an incomplete, piecemeal understanding of the modeling work. Authors dealt with the onslaught of papers in different ways. Some took a narrative approach rather than conducting a systematic review^8^ and few included preprints^9^. The number of papers included for analysis ranged from less than 20^10^ to 250^9^. Only a few papers explicitly stated their scoping process and selection criteria^9,11,12^. All of the reviews identified by our search are limited to papers published before August 2020^10–13^ or in 2020^9,14^. Many of these reviews are focused on model objectives and methodology^12–14^. This paper will provide a systematic review that covers papers up until August 20, 2021, with a focus on factors that are important for model utility, which we define as the capability of a model to provide insight for decision-makers and the general public.

This paper will analyze the inputs, outputs, and evaluation of predictive COVID-19 models, with a focus on elements that have been neglected in the current literature: data, uncertainty, performance evaluation, and limitations. We provide a unique quantitative evaluation of each of these elements, which enables stronger and more justified conclusions about trends and areas in need of improvement, with respect to modeling COVID-19 and future pandemics. In addition, we provide commentary on the elements of modeling that are not consistently covered in the literature but are crucial for building useful models: data infrastructure, information sharing systems, and model translation.

## Methods: Search Strategy and Selection Criteria

For the purposes of this literature review, we classified COVID-19 disease spread modeling into two major categories: retrospective analysis and prospective modeling. Retrospective modeling, or backward-looking analysis, has been applied throughout the outbreak to explore a variety of key questions such as inferring basic epidemiological characteristics like *R*_0_, incubation period, and fatality rate, reveal factors driving transmission, and assess the effectiveness of different interventions^15–17^. In contrast, prospective modeling is forward looking, and includes forecasts, projections, and future scenario analysis. Forecasting aims to predict near term epidemiological dynamics, often relying on data-driven methods and assuming that there will be minimal changes during the forecast period, while projections span over a much longer future time window, and thus must make assumptions about how the factors driving COVID-19 will change in the future. Scenario analyses produce multiple projections that explore the impacts of different sets of assumptions that vary factors like transmission rates and interventions.

Due to the substantial number of past and ongoing COVID-19 modeling efforts, we imposed significant constraints on the scope of this review to enable us to conduct a systematic, quantitative, and timely assessment of the relevant literature. Specifically, the following inclusion criteria defined our review scope:

1. Prospective modeling work on population-level dynamics of COVID-19: we include papers that provide future predictions for a specific location, including forecasting, projections, and future scenario analysis. We exclude retrospective modeling studies. Papers that only fit a model without providing out-of-sample predictions were not included.
2. Data-driven: we broadly define this as papers that incorporate COVID-19 data into the setup or fitting of the model. Those which only use parameters from the literature or rely on data from other viruses were excluded.
3. Geographic restriction: we only included papers that implement forecasting or projections to US counties, states, or at the national level, which restricts our analysis to papers working with the same data issues and in a similar context.
4. Journal restriction: Only papers published in English-language journals ranked in the top 10% based on the Scopus’ CiteScore for their respective field were included. While we recognize this will exclude important work, this criterion enabled a systematic approach to reducing the set of papers to a manageable level, while still capturing a quality, representative sample of the literature from which broader conclusions can be drawn.

The Scopus query we developed based on these criteria is included in the Supplement.

To minimize the chance of our search missing relevant papers, we also searched PubMed with the equivalent query. *Figure 1* outlines our scoping process and shows the number of papers screened out at each step.

**Figure 1.**
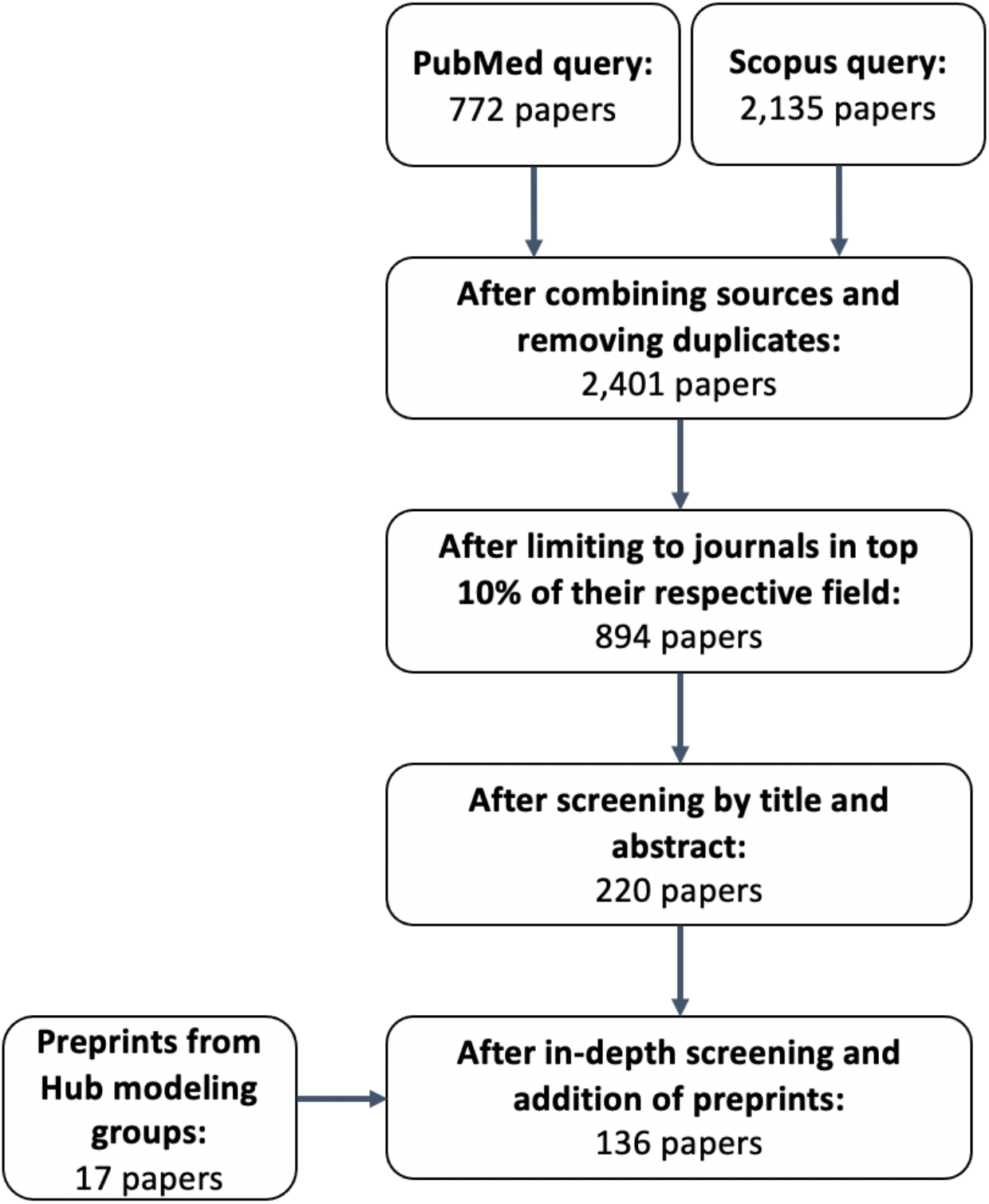
Scoping Process

The searches of Scopus and PubMed were carried out on August 20, 2021, and our final selection of papers was roughly evenly distributed from February 2020 to August 2021 (*Figure 2*). Notably, the top 10% criteria only reduced the number of papers to 37% of the original size, from 2,401 to 894 papers. After screening steps, we narrowed down to 119 published papers.

**Figure 2.**
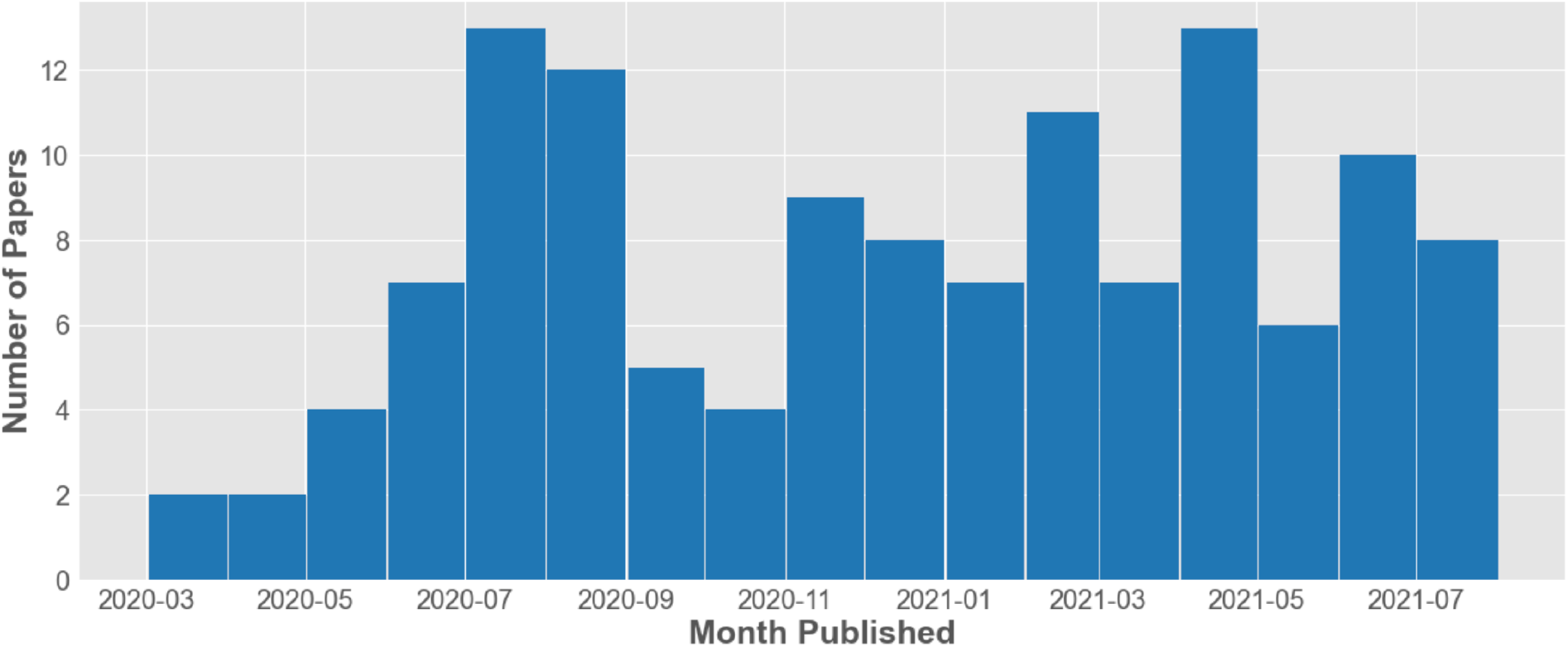
Number of Papers in our Analysis by Month of Publication.

We additionally considered preprints from authors known to be engaged in real-time modeling work, in an attempt to correct for the fact that many of the modeling groups most engaged in real-time work have not had the time to publish yet. We included preprints from modelers participating in the US COVID-19 Forecast Hub^2^. We also wanted to include the Scenario Modeling Hub^18–20^, but we did not find any preprints that met our criteria. While these papers do not have the validation that comes with peer-review, these models were used in real-time by a national public health agency, which we believe justifies their inclusion in this analysis. We found 17 preprints in the metadata provided by the modeling teams contributing to the Forecast Hub. Thus, 136 papers in total are included in our analysis. Despite our efforts, we acknowledge that we may still miss a portion of the COVID-19 modeling work that exists on preprint servers and on the websites of modeling groups.

We have designed our scoping process to obtain an objective and representative sample of the most recently published work, in order to draw larger conclusions about prospective COVID-19 modeling and highlight areas for improvement.

## Results

To conduct a quantitative analysis on the substance and quality of these studies, for each paper in our final set of papers we classified the following features: forecasting window, methodology, prediction target, datasets used, geographic resolution, quantitative uncertainty, performance evaluation, and stated limitations. We acknowledge that some of these categorizations are subjective and difficult to consistently extract from papers, especially the performance evaluation and stated limitations category. Thus, we narrowly define our categories and transparently discuss these definitions in this section. All of our categorizations were checked multiple times, and while we acknowledge that some of the categorizations we made could be disputed, we are confident that the overall conclusions still hold. The classification of papers for each category is shown in Supplementary Table 1^21–156^. *Figure 3* visualizes the relative size of each category and the most common connections between categories. Each line through the figure represents the categorizations of a single paper, so the thicker the line between two categories, the more often papers tend to fall into both of those categories.

**Figure 3.**
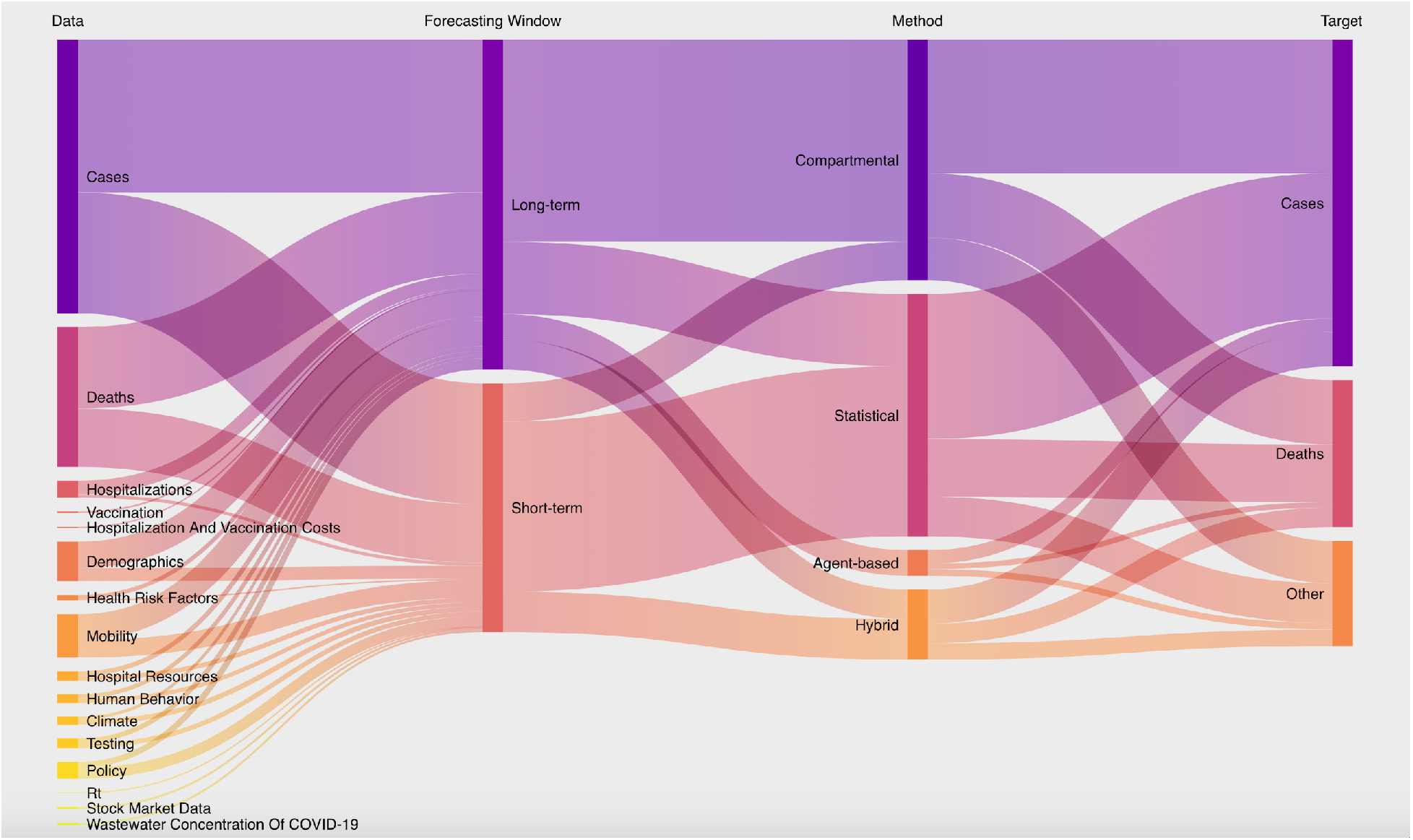
Sankey Diagram of the Connections Between Categorizations of our Analysis.

### Model Objective and Prediction Horizon

*Forecasts* are unconditional in the sense that they attempt to predict what will actually happen in the near future, while *projections* and *scenarios* are conditioned on the model’s assumptions about the future in order to extend the prediction horizon. We were unable to reliably categorize models into forecasts or projections due to inconsistent use of these terms and a lack of clear communication in papers on which purpose their model served. Therefore, as a proxy for model objective, we categorized papers into short-term predictions or long-term predictions, which broadly correspond to forecasts and projections, but not in every case. Since the COVID-19 Forecast Hub generates predictions for 1-4 weeks ahead, we chose four weeks as the cutoff for short-term predictions. Using this definition, 60% of papers made long-term predictions, and 46% of papers made short-term predictions. There were a small number of papers which produced both long-term and short-term predictions^30,72,84,92^. Note that because papers often fall into multiple categories, percentages in this analysis will not always add up to 100%. Within the category of papers conducting long-term projections, we also tagged papers with multiple scenarios, which provided multiple predictions based on different sets of assumptions. This could include modeling scenarios with different reopening speeds, non-pharmaceutical interventions, and vaccination rates. Of the 82 papers in the long-term projections category, 54 papers (66%) considered multiple scenarios.

### Methodology

Since many of the existing COVID-19 review papers go into more detail on this aspect of modeling^12–14^, we opted not to cover the methodologies beyond classification into three broad categories: compartmental models (SIR and variations), statistical models (machine learning, deep learning, ARIMA, etc.), and hybrid (a combination of compartmental and statistical models). We adopted a stringent definition of a hybrid model, requiring both compartmental and statistical layers of the model which go beyond using statistical approaches to fit parameters for a compartmental model. In addition, we noted if models used agent-based methods. We found that 47% of papers used a compartmental model, 43% used a statistical model, 12% used a hybrid model, and 9% used agent-based methods. There were a few papers which showed both a compartmental model and a statistical model^30,72,84,92^.

The wide interest in COVID-19 modeling from a variety of fields led to a diversity of methodologies being used in the literature. While a wider array of models can help provide more robust predictions, it is important for the appropriate methodology to be selected for the intended research objective. Especially for an ongoing public health crisis, modelers need to exercise extra care in providing model transparency and guiding readers towards an appropriate interpretation^157^, since their work can directly impact individual and group decisions.

### Target Variables

The most common target prediction variables were cases (89%), deaths (52%), hospitalizations (10%), and *R*_*t*_ (9%). Some of the lesser used target variables included growth rate, peak cases, and ICU admissions. 38% of papers had only one target variable, 43% of papers had two target variables, and 19% had more than two.

The target prediction variables were dominated by absolute numbers of cases and deaths, which aligns with the goals of the US COVID-19 Forecast Hub. Despite the continued desire for these targets from across the field of public health, government, industry, and the public, accurate prediction of them remains challenging^1^.

### Data Categories

Next, we quantified the categories of input data used to inform models. *Table 2* shows how we defined the data categories, including an in-depth look at the datasets used by papers in our analysis that attempt to capture COVID-19 behaviors.

**Table 2.** Data Categories.

| Data Category | Description | Examples |
| --- | --- | --- |
| Cases, Deaths | Epidemiological data on the number of cases or deaths and corresponding metrics. | Daily cases/deaths, cumulative cases/deaths, reproduction number, growth rate |
| Hospitalizations | Data related to hospitalization of COVID-19 patients. | Daily hospitalizations, active hospitalizations, ICU occupancy, hospital capacity |
| Testing | Data pertaining to COVID-19 testing in a population or location. | Daily tests, test positivity rate |
| Climate | Data describing the climate or any meteorological variables pertaining to a specific location, timeseries or static. | Daily precipitation, daily temperature, average temperature |
| Demographic | Demographic or socio-demographic information about the population of a specific location. | Population, age, race, income, rural/urban ratio |
| Health Risk Factors | Data which quantifies the health risk factors of the population in the context of COVID-19. | Prevalence of comorbidities, use of preventative services (doctor visits) |
| Mobility | Data which quantifies the movement of a population. | Google Mobility Trends (residential, grocery & pharmacy stores, parks, retail & recreation, workplaces, transit stations)<sup>158</sup><br>Unacast social distancing scoreboard (average mobility, nonessential visits, encounters density)<sup>159</sup><br>SafeGraph (trip counts at a census block group resolution)<sup>160</sup><br>Apple Mobility Trends (trends in apple maps routing requests)<sup>161</sup><br>Facebook Movement Range Maps (change in movement compared to baseline percent of population that stays home)<sup>162</sup><br>Flight data |
| Human Behavior | Data which quantifies the behavior or beliefs of a population in the context of COVID-19, excluding data on the mobility of a population. | Google search trends<sup>163</sup><br>Mask use per capita<sup>164</sup><br>Facebook's COVID-19 Trends and Impact Survey (timeseries of self-reported mask use and other social distancing behaviors)<sup>165</sup><br>New York Times Mask-Wearing Survey data (static)<sup>166</sup><br>Sentiment index constructed from COVID-19 news<sup>69</sup> |
| Policy | Data pertaining to policies relating to COVID-19. | Oxford COVID-19 Government Response Tracker (ordinal scale on stringency of many types of COVID-19 policies, including containment and closure policies, economic policies, health system policies, and vaccination policies)<sup>167</sup><br>State Level Social Distancing Policies: tracks dates and details of policies including emergency declarations, gathering restrictions, closures, stay-at-home orders, travel restrictions, isolation orders, and mask mandates<sup>168</sup> |

The most frequently used data categories were cases, deaths, mobility, demographics, and hospitalizations. 20% of papers used only one category of data, 39% of papers used two categories, 16% used three categories, and 25% used four or more categories *(Table 3)*.

**Table 3.**
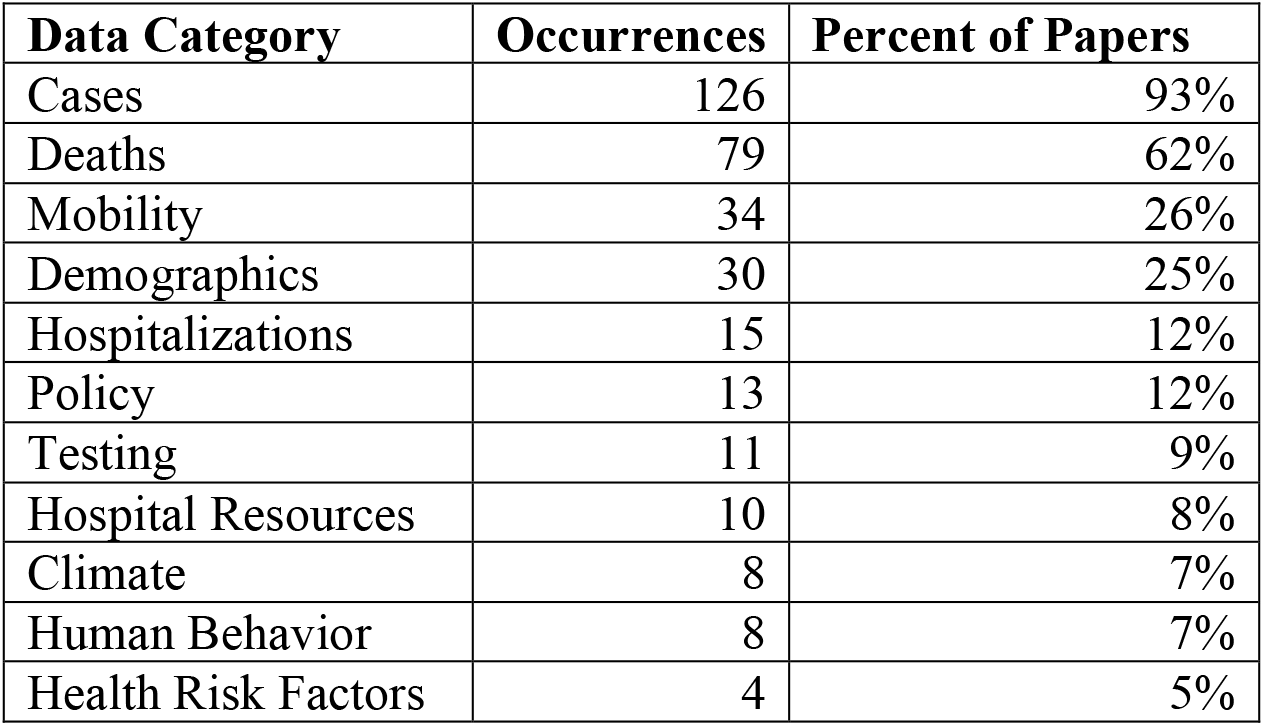
Top 10 Data Categories.

| **Data Category** | **Occurrences** | **Percent of Papers** |
| --- | --- | --- |
| Cases | 126 | 93% |
| Deaths | 79 | 62% |
| Mobility | 34 | 26% |
| Demographics | 30 | 25% |
| Hospitalizations | 15 | 12% |
| Policy | 13 | 12% |
| Testing | 11 | 9% |
| Hospital Resources | 10 | 8% |
| Climate | 8 | 7% |
| Human Behavior | 8 | 7% |
| Health Risk Factors | 4 | 5% |

The data sources informing predictions in our analysis were dominated by case and death data. Data used in 2 or less papers include vaccination, *R*_*t*_, wastewater surveillance, and economic data. Most modelers did not use non-epidemiological data sources, with mobility and demographic data as the main exceptions. The models that did use other data sources tended to incorporate a large number and variety of input data. We suspect that human behavior in response to COVID-19 could have an important impact on epidemiological dynamics, but the datasets currently available to capture this are limited. Mobility data can only capture a limited aspect of risk reduction behavior, and surveys of self-reported social distancing behaviors suffer from substantial sampling biases and low spatial and temporal resolution. Some other factors have been shown to be associated with COVID-19 dynamics, such as demographics, health risk factors, and climate, although little research has been done to rigorously test for whether these factors can improve predictive performance. These factors were rarely used in our sample of papers. Of particular interest due to the increasing impact of new variants on epidemiological dynamics, none of the papers in our sample utilized variant prevalence data. In the US, this data suffers from low sample size, sampling bias, and is difficult to use as a signal for predictive modeling.

### Geographic Resolution

We noted the geographic scale at which predictions were made, categorizing papers as national, state, or county-level and lower. 54% out of 136 of papers included a national level prediction, 36% at the state-level, and 34% at the county-level or smaller scale. Half of the models in our analysis were at the national level, which tends to be the easiest resolution to predict and the least useful for decision-making, which must often occur at the local level.

### Uncertainty

We analyzed which papers included a quantitative expression of uncertainty of their predictions, excluding those which only did so for model parameters. We found that half of papers (50% out of 136) did not express any uncertainty. 49% of papers included some form of confidence or prediction intervals. A sensitivity analysis was performed in 13% of papers^32,68^.

Half of the papers studied did not express any quantitative uncertainty around the forecasts, despite the highly uncertain and consequential nature of COVID-19 dynamics. The utility of forecasts for decision-makers depends on clear communication of uncertainty^169^, especially since point estimate predictions will rarely match ground truth data. Well calibrated expressions of uncertainty help stakeholders assess future risk and decide how to respond. For example, the difference between a 1% chance of exceeding hospital capacity versus a 25% chance could determine whether certain preparatory actions are taken. Additionally, expressing uncertainty is especially important to prevent harmful, incorrect interpretations of COVID-19 models. Clearly communicating uncertainty around predictions weakens the ability of actors to use a study in a misleading way to support their preexisting agenda.

### Performance Evaluation

We categorized the type of performance evaluation used for each model. We chose to conduct this analysis only for the subset of papers implementing short-term prediction models, which can be fairly evaluated against truth data. In contrast, the purpose of long-term projections is to compare multiple plausible scenarios of the future, not to predict what will actually happen. Therefore, evaluating the performance of these predictions is not possible using standard error metrics, since these models make assumptions about the future that do not match reality. For timeseries forecasts, the setup of train and test data should be representative of real-time forecasting conditions. Since the utility of a model is based on its ability to predict future dynamics, randomly excluded “out-of-sample” evaluation methods do not adequately describe performance. Instead, models should be trained using data up until a certain cutoff date and evaluated on data after that date. This preserves the fundamental challenge of forecasting: not knowing future data or trends. Within the subset of short-term studies considered, 75% of papers used some sort of performance evaluation metric to compare future-blind, out-of-sample predictions to ground truth data. The most commonly used metrics were mean absolute error, root mean square error, mean absolute percentage error, *R*^2^, mean square error, and coverage rate of prediction intervals. Out of the papers that did conduct a metric-based evaluation, only 13% evaluated the accuracy of confidence intervals. Within the group of 47 papers which conducted out-of-sample performance evaluation, 34% evaluated only one model, 55% compared performance metrics across multiple internal models, and 19% compared the performance metrics of their model against those of other models in the COVID-19 Forecast Hub *(Table 4)*. 15% of evaluated models used a baseline model for comparison.

**Table 4.** Categorization of Performance Evaluation for Short-Term Predictions

| Prediction Horizon | Performance Evaluation Category | Performance Evaluation Sub-Category |
| --- | --- | --- |
| Short-Term Predictions | Metric-Based Performance Evaluation – **75%** | Evaluate One Model – **34%** |
|  |  | Evaluate Multiple Internal Models – **55%** |
|  |  | Evaluate and Compare to Models from Forecast Hub – **19%** |
|  | No Performance Evaluation – **25%** | N/A |

Most modelers (75%) quantified the performance of their model relative to truth data, but most did not evaluate their model on predictions made across a timespan that included varying epidemiological dynamics. In order to quantify this, we counted how many dates papers showed predictions **from**. For example, let’s consider a paper that shows a model prediction using data up until September 1^st^ and predicts future case counts on the 8^th^, 15^th^, 22^nd^, and 29^th^. This would be a prediction made from a single date. If this paper adds another prediction made from (model uses data up until) October 1^st^ and predicts weekly values for the next 4 weeks, this paper would be showing predictions made from two dates, which cover a month-long timespan (September 1^st^ to October 1^st^). We defined the category this way in order to make sure we could reliably extract this data from each paper. Our analysis found that among short-term models, more than half (55%) only showed a prediction made from a single date. 28% of papers showed predictions made from multiple dates over a timespan that was less than 2 months long, while 17% covered a timespan longer than 2 months. From the COVID-19 Forecast Hub, we know that predictive accuracy of models varies widely over time, especially with respect to epidemiological trends^2^. Therefore, failing to evaluate a model in a variety of epidemiological dynamics severely limits the generalizability of performance evaluation and the ability to make fair comparisons between models. In addition, one-third of papers (34%) that completed a quantitative performance evaluation did not compare their model to a baseline or any other models, so it is unclear whether the model provides any improvement over a naïve model. The COVID-19 Forecast Hub uses a baseline model that assumes no change in incidence over the next four weeks. According to historical error metrics calculated on September 8^th^, 2021, only 25% of models outperformed the baseline model for cases while 75% outperformed the baseline for deaths, calculated by relative mean absolute error and weighted interval score^170^. Thus, comparison to a baseline model provides context that provides important information about the utility of a model. Many papers did not cover the specific methodology of their performance evaluation, which limited our ability to provide more specific analyses in this review. Authors should clearly state the dates of the training period, the dates predictions were made from, how error metrics were computed and aggregated, and whether metrics are computed in-sample or out-of-sample. In addition, models that aim to contribute to real-time forecasting efforts should use input data as it was available at the date predictions are made from. Without thorough performance evaluation, the broader scientific community will be unable to determine which approaches are working and build knowledge on best practices.

### Model Limitations

Authors stated six main categories of limitations: disregarded factors (39%), data quality (28%), unknowable factors (26%), limitations specific to the methods used (22%), data availability (16%), and limited generalizability (8%). We define unknowable factors as those that cannot be known at the time predictions were made, like future implementation of non-pharmaceutical interventions or the emergence of new variants during the prediction horizon. In contrast, disregarded factors have some relevant data or information available at the time of the analysis, but the authors choose to disregard it for simplicity, like the demographic breakdown of populations or healthcare capacity of different regions. A third of the papers in our analysis (36%) did not list any limitations in an accessible section of the paper, which we considered to be in the discussion, conclusion, or in a separate section named limitations. In most cases, all of these types of limitations are relevant to COVID-19 models. In addition, our categorization does not give information about how thoroughly these limitations categories were discussed. For COVID-19 applications, clearly stating model limitations is crucial to help the public understand the appropriate way to interpret results.

### Multidisciplinary Nature of the COVID-19 Literature

The highly consequential nature of the COVID-19 pandemic has attracted modeling experts from a variety of different fields. The top five journal subject areas represented in our final set of papers, in order from most to least frequent, are applied mathematics, multidisciplinary, general physics and astronomy, general mathematics, and statistical and nonlinear physics. Notably, public health did not appear in the top five subject areas. Our final set of papers represented 52 journals. The most common journals were *Chaos, Solitons, and Fractals, PLOS One*, and *Scientific Reports* (*Figure 4*). We were unable to conduct a thorough analysis on the contributions to COVID-19 modeling from different fields due to the difficulty of classifying papers into different disciplines based on their journal and the inherent interdisciplinarity of the work. However, we completed sub analyses on the group of papers from Forecast Hub modelers and a physics and math journal with interesting characteristics.

**Figure 4.**
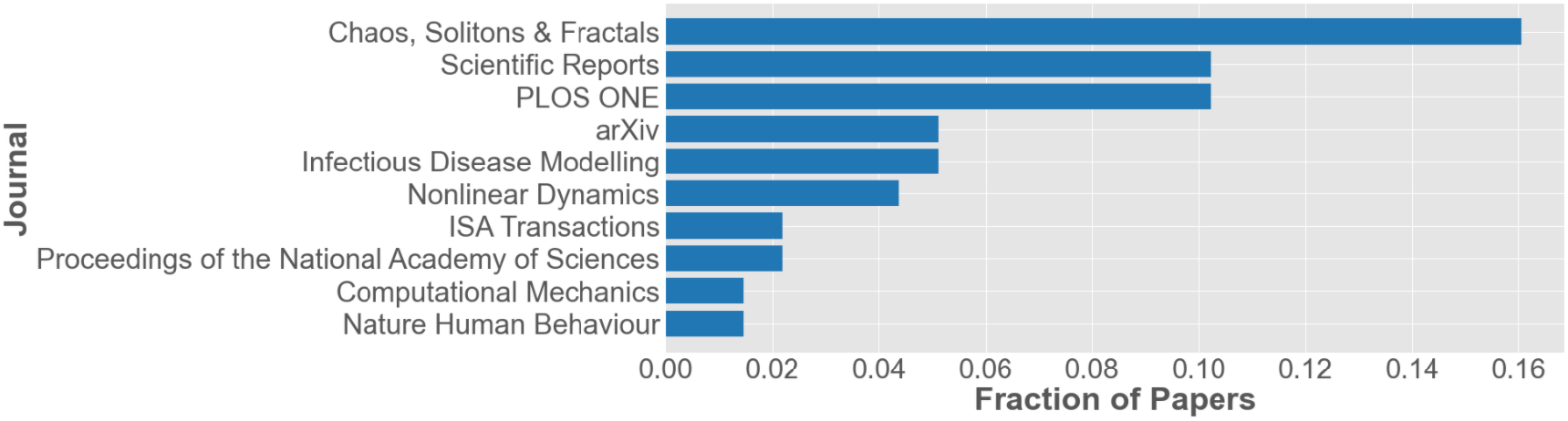
Top 10 Journals in the final set

Papers from the journal *Chaos, Solitons, and Fractals*, which specializes in mathematics and physics, composed a large portion of our final set of papers (22 out of 118 published papers)^26,51,60,64,66,73,75,77–81,83,85,87,100,102,103,118,124–126^. So, we conducted a separate analysis of this subset, shown in *Table 5*. Notably, this journal was heavily skewed towards national level predictions (95%) and favored statistical models (77%) and short-term predictions (64%). Papers from this journal were significantly less likely to express quantitative uncertainty compared to our sample of all papers (14% versus 50% for all papers). Out of short-term models only, *Chaos* papers were less likely to do a performance evaluation that compares predictions to ground truth data (64% versus 75% for all papers), and significantly more likely to only make predictions from a single date (93% versus 55% for all papers). In addition, papers from this journal were significantly less likely to state limitations (23% versus 64% for all papers).

**Table 5.** Comparison of All Papers, Forecast Hub Preprints, and Chaos, Solitons, & Fractals

| Categories | All Papers<br>(N=136) | *Chaos,*<br>*Solitons, &*<br>*Fractals*<br>(N=22) | Forecast Hub<br>Papers and<br>Preprints<br>(N=20) |
| --- | --- | --- | --- |
| Prediction Horizon |  |  |  |
| Short-term Predictions | 46% | 64% | 70% |
| Long-term Predictions | 60% | 45% | 40% |
| Methodology |  |  |  |
| Compartmental | 47% | 18% | 35% |
| Statistical | 43% | 77% | 45% |
| Hybrid | 12% | 5% | 20% |
| Agent-based | 9% | 0% | 5% |
| Geographic Level |  |  |  |
| National | 54% | 95% | 25% |
| State | 36% | 14% | 65% |
| County or Smaller | 34% | 5% | 55% |
| Uncertainty |  |  |  |
| Expressed Quantitative Uncertainty | 50% | 14% | 55% |
| Sensitivity Analysis | 13% | 0% | 5% |
| Performance Evaluation |  |  |  |
| Comparison to Ground Truth (out of<br>short-term models only) | 75% | 64% | 86% |
| Only Made Predictions from One Date | 55%<br>17% | 93%<br>7% | 7%<br>50% |
| Made Multiple Predictions over a Timespan greater than 2 months |  |  |  |
| Limitations |  |  |  |
| Authors discussed limitations | 64% | 23% | 65% |

We also conducted an analysis on papers written by authors that contributed to the COVID-19 Forecast Hub, which includes 17 preprints^138–140,142–150,152–156^ and 3 published papers^137,141,151^. 70% of these papers made short-term predictions and 40% of these papers made long-term predictions. Although these papers were cited by teams in the metadata of their submissions to the COVID-19 Forecast Hub, which focuses on one to four week predictions, these preprints are not necessarily on the exact model and application that was submitted to the Forecast Hub. Despite being mostly preprints with many serving to provide a brief explanation a model being used in real-time, these papers were more likely to express uncertainty, have forecasts for state and county levels, and conduct performance evaluation than the full set of papers. In addition, Forecast Hub papers were significantly more likely to show and evaluate predictions made from several dates over a timespan greater than 2 months (50% versus 17% for all papers). A significant advantage of the hub approach is that it encourages good practices in terms of uncertainty, evaluation, and high geographic resolution. Additionally, the real-time sharing of forecasts ensures that predictions were truly future-blind.

### Epidemic Information Systems

Most of the operational models that have been used in real-time to inform decision-making have not yet made it into the published literature. Although we tried to correct for this by adding some preprints from the COVID-19 Forecast Hub, the combination of hub preprints and papers only made up a small fraction of the papers in our analysis (15% of 136 papers). The difficulty of scoping this literature review to capture real-time work highlights that the current publication system is not suited to the rapid information-sharing needs of outbreaks, even in the context of increased use of preprints^171^. As COVID-19 research took off, editors were overwhelmed with so many submissions that the already slow process of peer-review by epidemic standards was further slowed, forcing more work onto preprint servers. Preprints excel at quickly sharing new research but lack the quality assurance provided by peer-review—a particularly important aspect for models used to inform urgent decisions. In addition, the ability to continuously update models is incredibly important for modeling a rapidly evolving pandemic. This need for continuous updating of operational models means that real-time modelers have little time to spend on making sure their documentation is current and available to other researchers. We need an information sharing system that is better suited to the needs of outbreaks, which strikes a balance between speed and quality, expects updates, and can fit into the busy schedule of real-time modelers. For example, the 2019 Novel Coronavirus Research Compendium^172^ curates and assesses recent research papers that are receiving a lot of attention, serving as a faster, bare-bones version of peer review.

The lack of an efficient system for tracking COVID-19 research increases the difficulty of and limits the insights gained from literature reviews. We had to design an intensive scoping process, which narrowed the work to a subset of prospective modeling. Other reviews adopted their own narrow scope, creating a body of COVID-19 modeling literature reviews which amount to a piecemeal, incomplete picture of the efforts of researchers.

Another aspect of the COVID-19 literature that limited the depth and insight of our analysis was a lack of standards for reporting on COVID-19 models. Papers did not consistently state the precise objective of their model (unconditional forecast or assumption-based projection), detail their methodology, express uncertainty, evaluate performance across a long, varied timespan, and clearly list their limitations. Without this information, our ability to synthesize insights from the research to determine best practices is limited. In response to these concerns, the EPIFORGE 2020 guidelines were developed and recommend consistent terminology, a clear definition of study purpose and model targets, identification of prospective versus retrospective work, comparison to a baseline model, a non-technical summary of results, and full documentation of: data sources, data availability, data processing, methods, assumptions, code, model validation, forecast accuracy evaluation, uncertainty, limitations, interpretation, and generalizability^157^. Consistent sharing of this information for epidemiological predictions would improve the consistency, reproducibility, comparability, and quality of epidemic forecasting reporting.

## Discussion: Commentary on Data Quality and Model Translation

The goal of this literature review was to cover the prospective COVID-19 modeling research that has been done, with a focus on model utility, and identify areas for improvement. Our analysis covered many topics that are central to the success of COVID-19 modeling, but there are some aspects of modeling that are not directly covered in the COVID-19 papers we analyzed but are crucial for the utility of our work, which we will comment on in this section.

### Data: Quality and Availability

Data quality is one of the most important drivers of model performance. If the data are inconsistent or not a true reflection of reality, models have no reliable ground truth to learn from. Unfortunately, the public health infrastructure in the US was not equipped to provide timely, high-quality COVID-19 data, necessitating several disparate efforts to provide more accessible COVID-19 data^173,174^. Despite the valiant efforts of data collectors, there are several flaws in the underlying reporting system that are difficult to account for in modeling. For example, the decision-making on how to collect COVID-19 data fell to the states, and each state had their own reporting idiosyncrasies, like defining what counts as a COVID-19 case and death, whether this includes probable cases/deaths, and how to define a “probable” case/death. Another frequent occurrence for case and death data is a large artificial spike or drop in the timeseries due to errors such as discovering old cases or deaths, correcting a past overreport, or a technical issue. When errors occur, many state public health dashboards did not clearly explain the cause of the anomaly in a timely, accessible fashion, leaving modelers to guess what the correct timeseries should be and whether it might be updated in the future. Other COVID-19 data, like counts of vaccinations, tests, hospitalizations, and prevalence of variants, suffer from their own data quality issues, mainly due to an incapable data reporting infrastructure, lack of universal data standards, and sampling bias. Number of tests is a particularly important factor due to its influence on number of reported cases.

These inconsistencies impinge upon modelers’ ability to provide accurate predictions. Modelers have been forced to wonder whether to aim to predict unreliable reported data or actual numbers of infections and deaths, which for COVID-19 will never be fully observed. If modelers aim for the latter, they must attempt to correct for the layers upon layers of issues that distance reported data from the truth. In order to give modelers the best chance of success, we need to build a data system that provides open, timely, standardized data at a high spatial and temporal resolution. Models built using existing indicators have not been able to capture inflection points, a reflection of the difficulty of predicting phenomena driven by human behavior. However, we currently do not have reliable data that measures COVID-19 risk reduction behaviors at a high temporal and spatial scale, so obtaining this data is a promising avenue for improving predictions. Another data source with potential is the prevalence of variants of concern, which could provide a warning sign of an upcoming wave. Therefore, a more robust genomic surveillance system could be a great resource for improving the predictive capability of models, especially in light of the impact of new variants on recent epidemiological dynamics.

Yet another factor that complicates the translation of relevant datasets into accurate models is that data availability, quality, and relationships between variables change throughout the outbreak. For example, mobility data was found to be associated with case growth during the first wave in the US, but not for later waves^175^. The introduction of vaccines, new variants, different kinds of COVID-19 tests, and policy changes have all played different roles in driving epidemic dynamics and the resulting risk reduction behaviors throughout the pandemic. This constant changing of dynamics that drive COVID-19 transmission coupled with data quality limitations has hindered model accuracy.

### Translation

#### Transparency

Although the translational aspect of modeling is often overlooked in academic papers, better communication of modeling is an integral part of producing useful models. One of the most important aspects of successful modeling translation is transparency in how models are built and how they should be used, which is covered in the aforementioned EPIFORGE 2020 guidelines^176^. Since COVID-19 modeling attracted many researchers without prior infectious disease modeling experience, the adoption of epidemic reporting guidelines is especially helpful to help modelers support decision-makers and avoid causing unintentional harm. To advance knowledge of best practices for translation, modelers should prioritize documentation of the process of sharing models with decision-makers when possible. For example, this paper documents the process of modelers working with policymakers in Utah to provide COVD-19 decision support^177^. By neglecting to share their experiences and knowledge on translation of models, modelers are missing an opportunity to harness the collaborative power of academia to identify best practices for translation and boost the utility of their work.

#### Control the Messaging

Due to the uncertainty and fear surrounding an unprecedented outbreak, modeling results were sometimes sensationalized by the media or were bent to serve a predetermined political purpose. To prevent misrepresentation, modelers must be explicit in stating how the assumptions and limitations shape the appropriate interpretation and control the corresponding public health messaging. Most of the papers in our analysis did not discuss what questions the model is appropriate to answer and how results should be applied to decision-making. Due to the serious consequences of misunderstandings, modelers have a responsibility to facilitate appropriate interpretation of our work. Many of these misunderstandings are rooted in the public lacking basic understanding of models and how they should be used. Therefore, a concerted effort from the modeling community is needed to build public understanding of the basic principles of model translation. For decision-makers, the best approach is often direct collaboration with modelers, since modeling and its interpretations are often complicated. These mutually beneficial relationships allow modelers to better understand the needs of decision-makers and help decision-makers better understand the nuances of epidemic modeling.

One aspect of modeling that could be better designed for easy interpretation is the selection of prediction targets. In the Forecast Hub, dozens of modeling groups have predicted the absolute number of cases, deaths, and hospitalizations for the next four weeks throughout the COVID-19 pandemic. The ensemble, which is consistently one of the top performing models, has shown poor performance during times of rapid changes in trends^1^, which are the most crucial moments for decision-making. This is especially pronounced in forecasts of cases and hospitalizations and less so for deaths. In response, modelers should consider new targets that may be more aligned with what models are able to accurately predict. For example, categorical predictions that classify a region into rapid growth, moderate growth, flat, moderate decline, or rapid decline, still convey the information that most model users want to know but is more interpretable for the average person. Since numerical forecasts of the precise number of cases or hospitalizations several weeks in the future have not been reliable over the course of the first 18 months of the pandemic, trying new experimental targets at different spatial and temporal scales could help provide models with more feasible targets. This could in turn enhance public trust in modeling. In addition, modelers tackling a broader range of targets can provide a more complete picture of relevant outcomes for stakeholders.

A crucial aspect of model translation is communicating that there are a range of plausible outcomes and that point predictions should not be interpreted as what will actually happen. Thus, modelers must clearly communicate uncertainty, but the way that modelers currently do so can be confusing for the public. For example, the 50% and 95% confidence intervals adopted by the COVID-19 Forecast Hub are often wide, spanning both upward and downward trends, which can be difficult to interpret. Therefore, modelers need to explore alternative ways to communicate uncertainty that translate statistical concepts into formats that are more accessible to the public, like providing the percent chance that the trend will be increasing, flat, or decreasing. More clarity in this aspect of communication will build public trust in modeling and help to prevent actors with ulterior motives from using a paper to support their preconceived agenda.

#### Value of Translational Work

A barrier to modelers in academia engaging in critical but time-consuming translational work is that incentive structures usually reward publishing papers and other traditional forms of academic achievement. However, most of the crucial work that was done for this pandemic was in building, updating, and communicating models and their results in real-time. In order for our research to maximize its impact in mitigating outbreaks, we need to recognize and elevate the value of translational work.

## Conclusion

This analysis examined a subset of the COVID-19 modeling literature, focused on data-driven, prospective modeling, and identified several opportunities to improve the utility of outbreak modeling. In response to significant scoping challenges, we selected a sample that should represent the best modeling papers and still found them to be substantially lacking in some of the areas that are most crucial for translating models into useful insight for decision-makers and the general public. As of August 20, 2021, we found that most of the translational modeling efforts that were (and in some cases still are) operational during the COVID-19 pandemic had not found their way into the peer-reviewed literature.

The main takeaways of this literature review relate to adopting epidemic forecasting standards, investing in quality datasets, engaging in thoughtful model translation, and creating a suitable information sharing system. Adopting the EPIFORGE 2020 guidelines address many of the issues identified in this review, including the need to be transparent about the methods, express uncertainty, thoroughly evaluate performance, state limitations, and discuss appropriate interpretations. Curating high-quality datasets, especially for ground truth epidemiological data, COVID-19 behaviors, and variant prevalence could greatly increase modelers’ chance of success. By thinking about translation during the modeling process, like when choosing prediction targets and uncertainty methods, modelers can make their model outputs more understandable for the public. Lastly, the creation of an information sharing system suited to the needs of an epidemic would allow faster advancement of knowledge and derive the maximum benefit from the hard work of COVID-19 modelers.

## Data Availability

All data produced in the present work are contained in the supplement.

## Contributors

LG, KN, and SJ contributed to the conceptualization and design of the study. KN, SJ, and FP collected the data and conducted the analysis. FP and SJ made the figures. KN led the writing of the original draft. NGR, KG, ECL, ST and LG edited the manuscript. LG supervised the study and acquired funding. KN and SJ have verified the underlying data. All authors had full access to the data and approved the manuscript for publication.

## Acknowledgements

KN, SJ, and LG were funded by the NSF Rapid Response Research grants, Award ID 2108526 and 2028604. NGR has been supported by the National Institutes of General Medical Sciences (R35GM119582). The content is solely the responsibility of the authors and does not necessarily represent the official views of NIGMS or the National Institutes of Health. KG and FP were funded by the SMDM Covid Modeling Accelerator.

## Supplementary Information

### Scopus Query

TITLE ({covid-19} OR {sars-cov-2})

AND TITLE-ABS-KEY (“model*” OR “forecast*” OR “project*” OR “predict*”)

AND TITLE-ABS-KEY (“US” OR “USA” OR “United States” OR “America”)

AND NOT TITLE ({protein} OR {clinical} OR {ct} OR {mental health} OR {psychological} OR {cell} OR {cellular} OR {rna} OR {diagnos*} OR {antiviral} OR {antibod*} OR {plasma})

AND NOT TITLE-ABS-KEY (“cancer” OR “molecul*” OR “cytokine” OR “receptor” OR “protein” OR “x-ray” OR “surgery” OR “surgical”)

AND (LIMIT-TO (PUBSTAGE, “final”)) AND (LIMIT-TO (DOCTYPE, “ar”)) AND (LIMIT-TO (LANGUAGE, “English”)).

**Supplementary Table 1.**
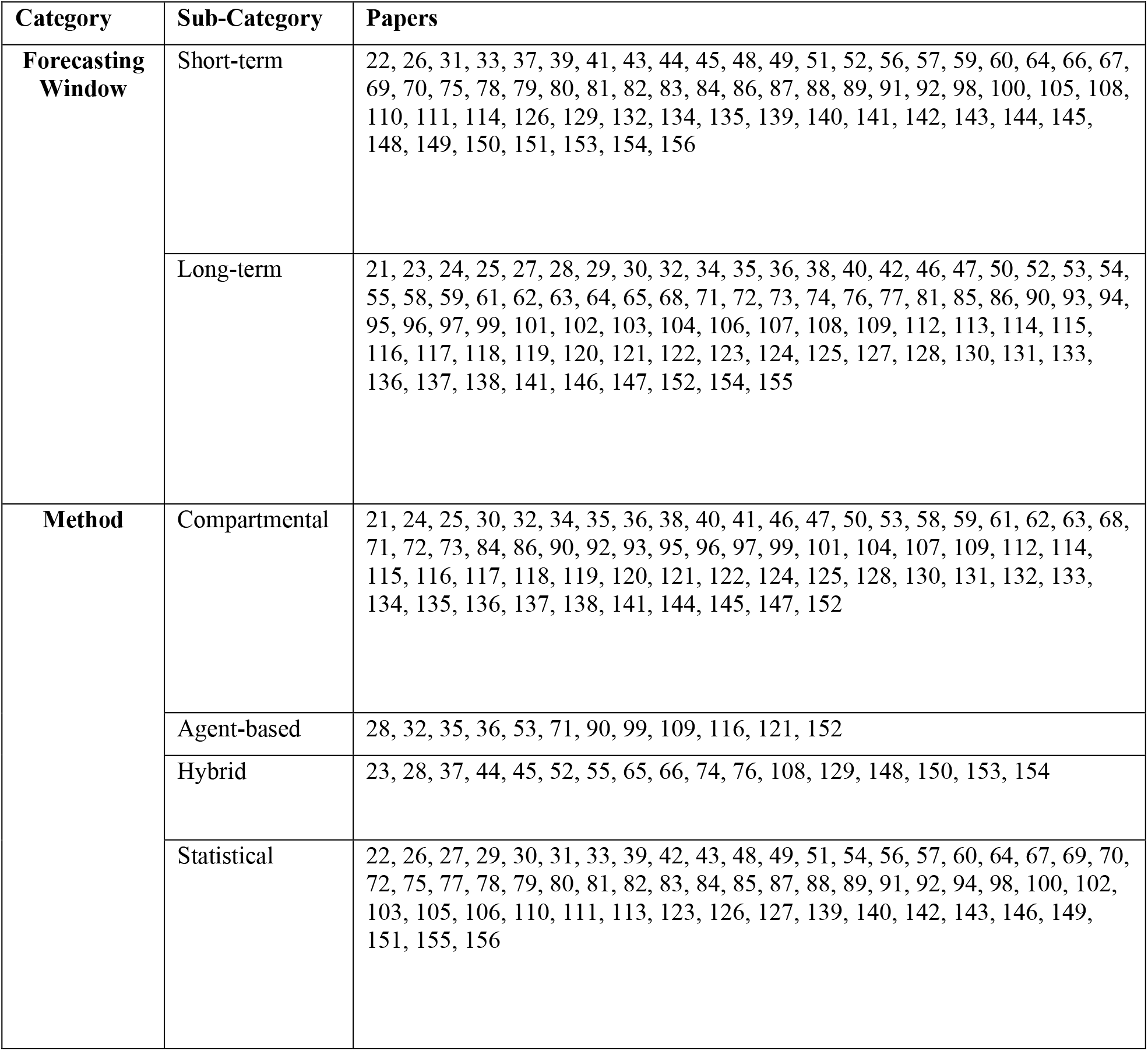
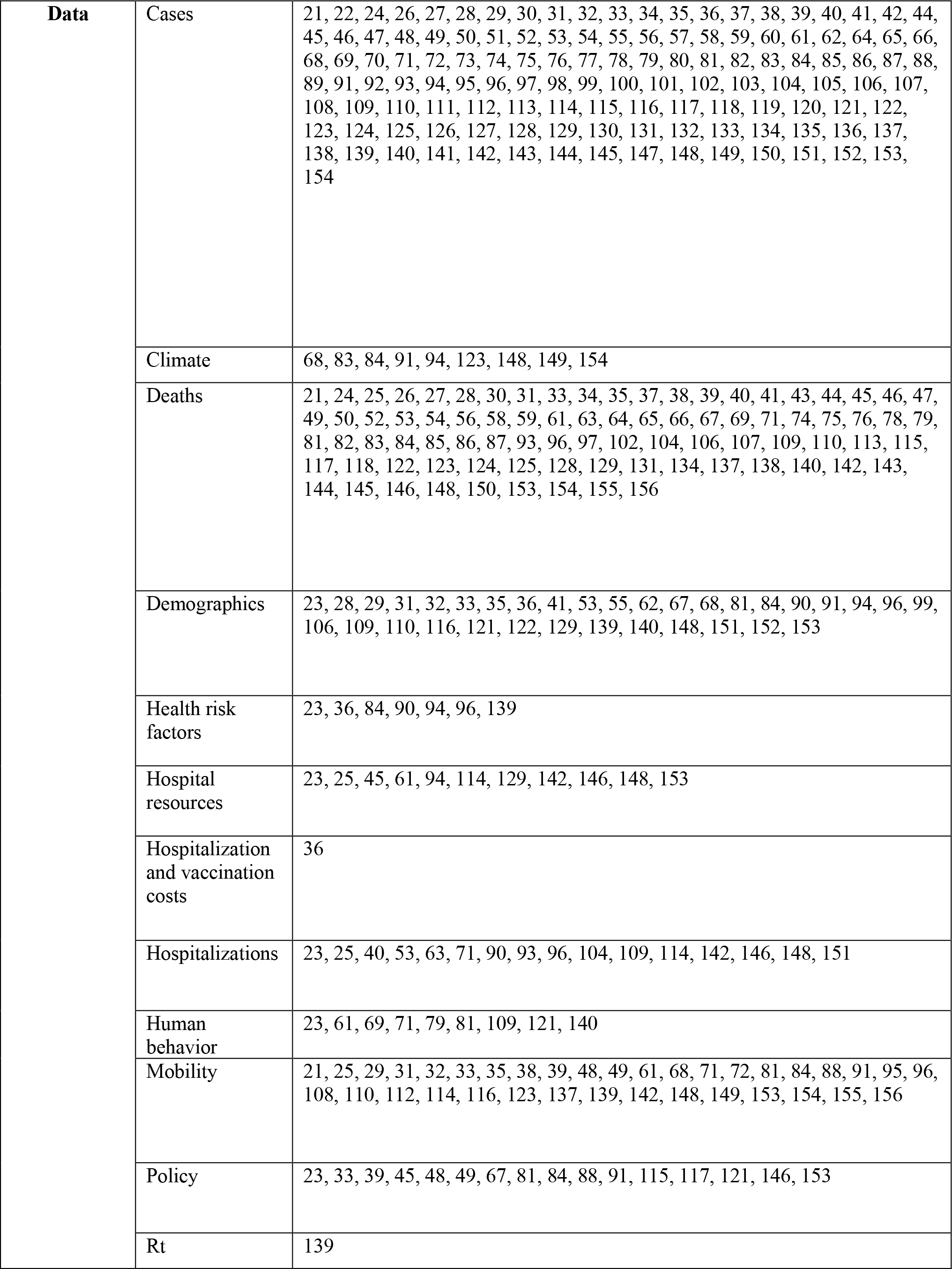
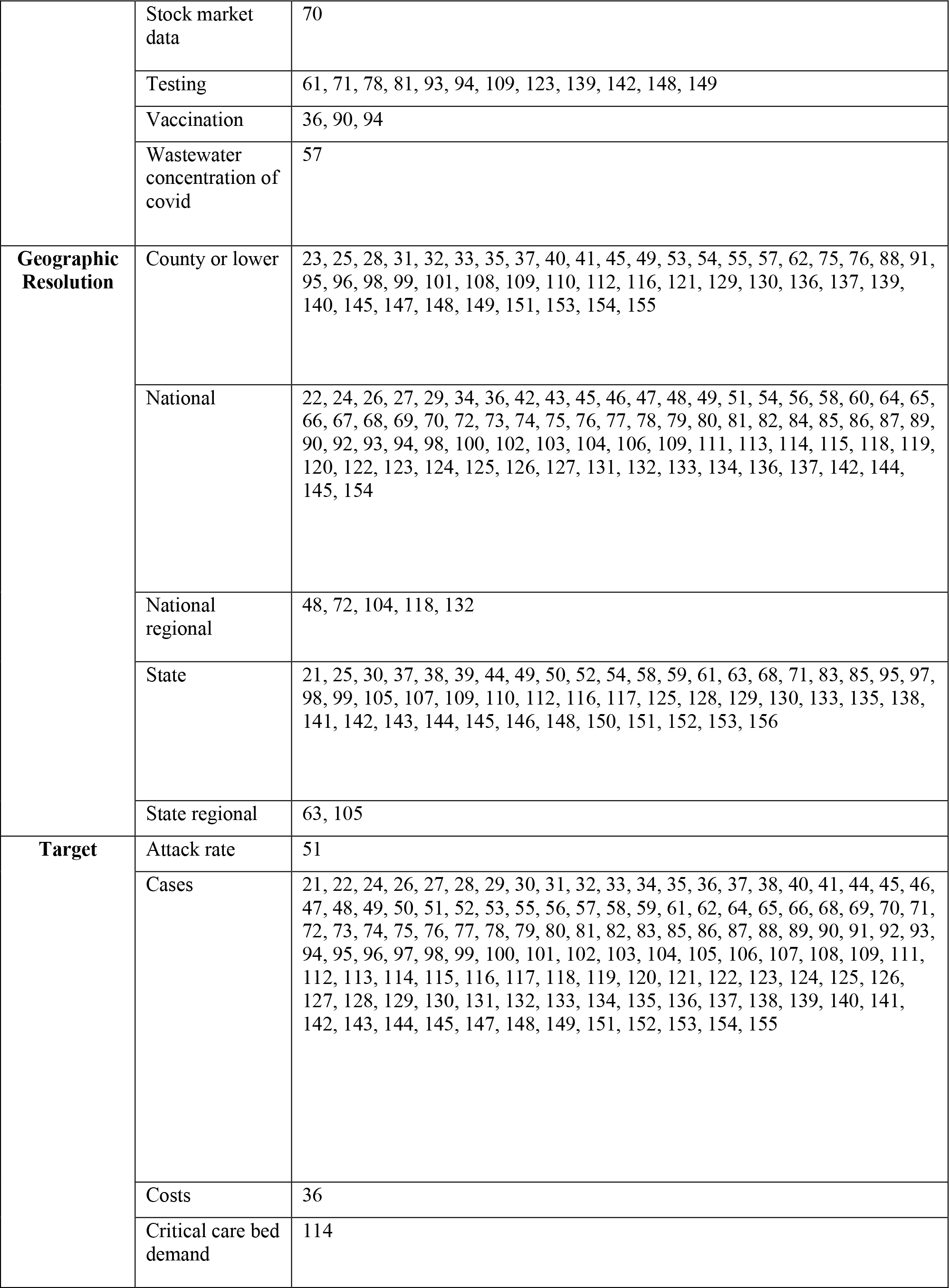
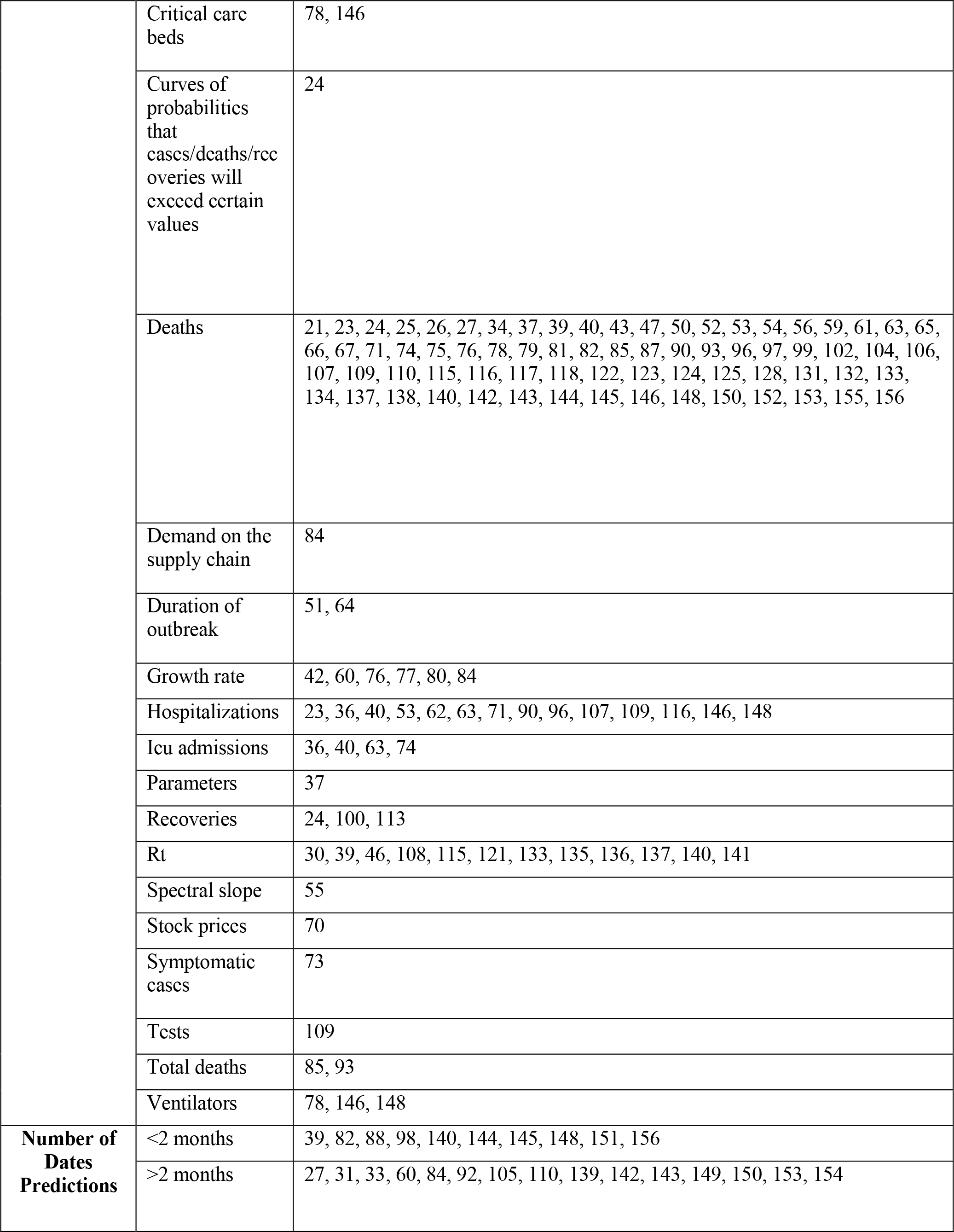
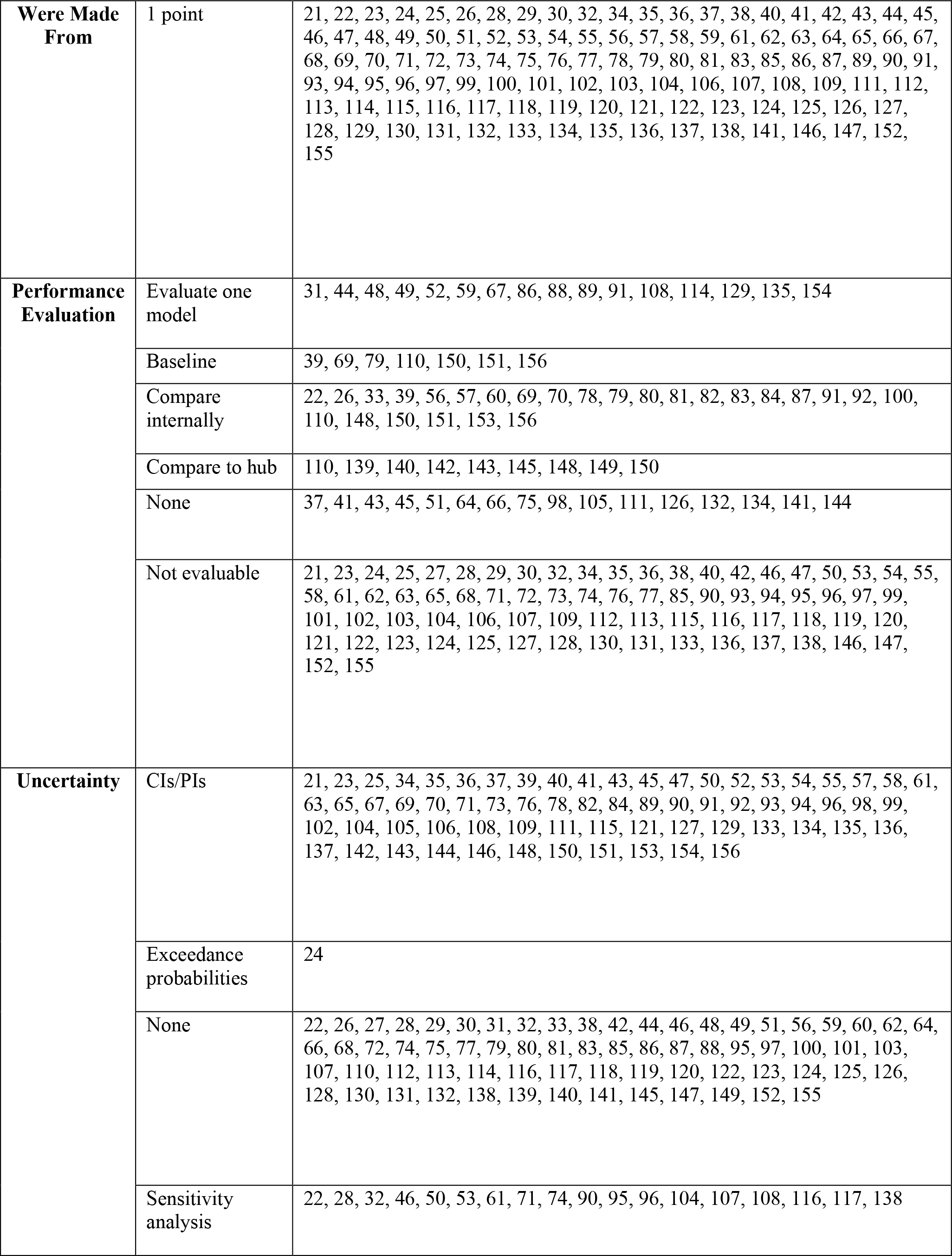
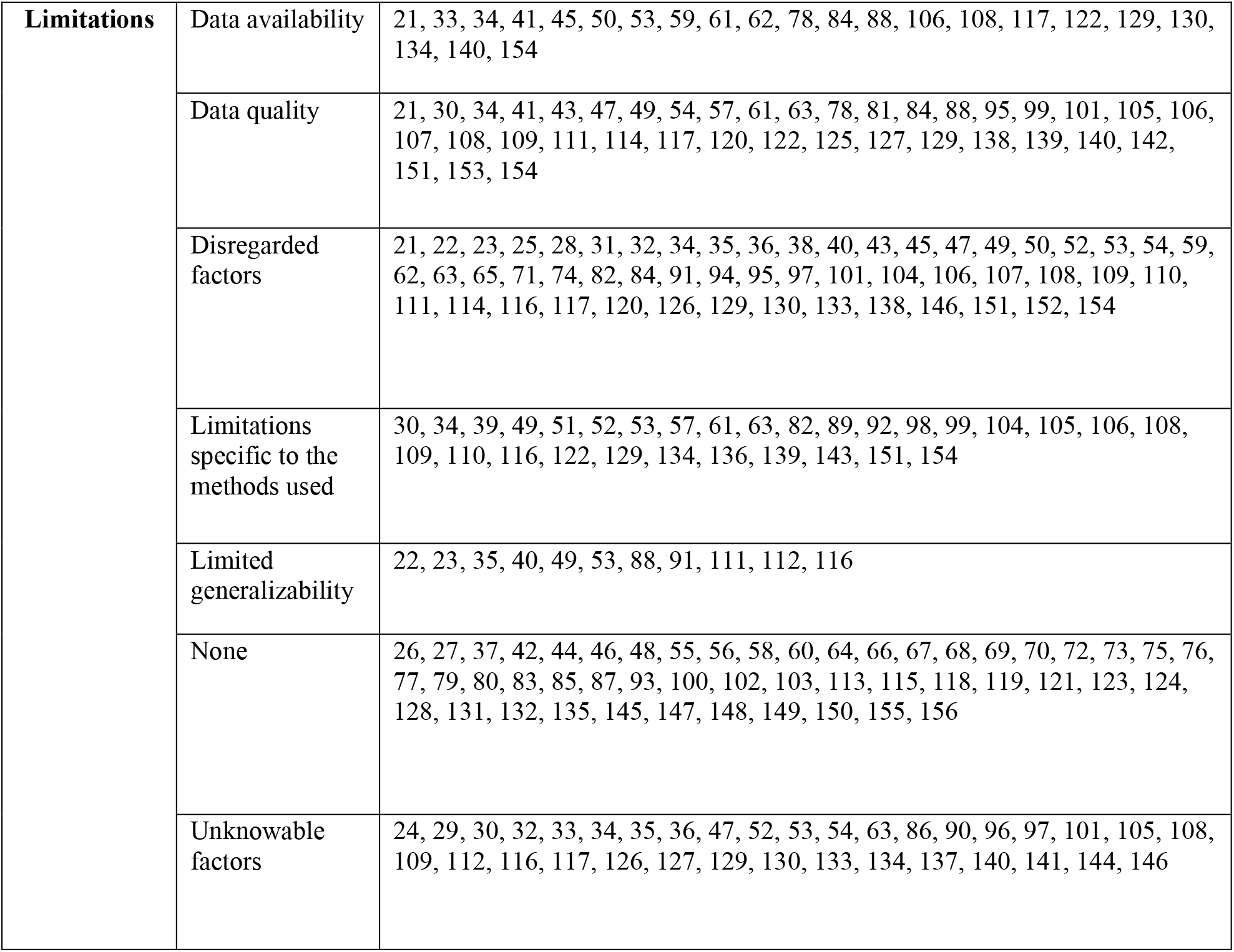
Categorizations for All Papers.

## References

1. On the predictability of COVID-19 - International Institute of Forecasters [Internet]. [cited 2021 Dec 6]. Available from: https://forecasters.org/blog/2021/09/28/on-the-predictability-of-covid-19/

2. Cramer EY, Ray EL, Lopez VK, Bracher J, Brennen A, Castro Rivadeneira AJ, et al. Evaluation of individual and ensemble probabilistic forecasts of COVID-19 mortality in the United States. Proceedings of the National Academy of Sciences [Internet]. 2022 Apr 12 [cited 2022 Apr 9];119(15). Available from: https://pnas.org/doi/full/10.1073/pnas.2113561119

3. Reich NG, Brooks LC, Fox SJ, Kandula S, McGowan CJ, Moore E, et al. A collaborative multiyear, multimodel assessment of seasonal influenza forecasting in the United States. Proceedings of the National Academy of Sciences [Internet]. 2019 Feb 19 [cited 2021 Dec 14];116(8):3146–54. Available from: https://www.pnas.org/content/116/8/3146

4. FluSight: Flu Forecasting | CDC [Internet]. [cited 2021 Dec 14]. Available from: https://www.cdc.gov/flu/weekly/flusight/index.html

5. James LP, Salomon JA, Buckee CO, Menzies NA. The Use and Misuse of Mathematical Modeling for Infectious Disease Policymaking: Lessons for the COVID-19 Pandemic: https://doi.org/101177/0272989X21990391 [Internet]. 2021 Feb 3 [cited 2021 Jul 11];41(4):379–85. Available from: https://journals.sagepub.com/doi/full/10.1177/0272989X21990391

6. Press WH, Levin RC. Modeling, post COVID-19. Science (1979) [Internet]. 2020 Nov 27 [cited 2021 Nov 20];370(6520):1015. Available from: https://www.science.org/doi/abs/10.1126/science.abf7914

7. Ioannidis JPA, Cripps S, Tanner MA. Forecasting for COVID-19 has failed. International Journal of Forecasting. 2020 Aug 25.

8. Adiga A, Dubhashi D, Lewis B, Marathe M, Venkatramanan S, Vullikanti A. Mathematical Models for COVID-19 Pandemic: A Comparative Analysis. Journal of the Indian Institute of Science 2020 100:4 [Internet]. 2020 Oct 30 [cited 2021 Jul 11];100(4):793–807. Available from: https://link.springer.com/article/10.1007/s41745-020-00200-6

9. Gnanvi JE, Salako KV, Kotanmi GB, Glèlè Kakaï R. On the reliability of predictions on Covid-19 dynamics: A systematic and critical review of modelling techniques. Infect Dis Model. 2021 Jan 1;6:258–72.

10. Zawadzki RS, Gong CL, Cho SK, Schnitzer JE, Zawadzki NK, Hay JW, et al. Where Do We Go From Here? A Framework for Using Susceptible-Infectious-Recovered Models for Policy Making in Emerging Infectious Diseases. Value in Health. 2021 Jul 1;24(7):917–24.

11. Shankar S, Mohakuda SS, Kumar A, Nazneen PS, Yadav AK, Chatterjee K, et al. Systematic review of predictive mathematical models of COVID-19 epidemic. Medical Journal Armed Forces India. 2021 Jul 1;77:S385–92.

12. Xiang Y, Jia Y, Chen L, Guo L, Shu B, Long E. COVID-19 epidemic prediction and the impact of public health interventions: A review of COVID-19 epidemic models. Infect Dis Model. 2021 Jan 1;6:324–42.

13. Guan J, Wei Y, Zhao Y, Chen F. Modeling the transmission dynamics of COVID-19 epidemic: a systematic review. Journal of Biomedical Research [Internet]. 2020 Nov 1 [cited 2021 Nov 21];34(6):422. Available from: /pmc/articles/PMC7718076/

14. Rahimi I, Chen F, Gandomi AH. A review on COVID-19 forecasting models. Neural Computing and Applications [Internet]. 2021 Feb 4 [cited 2021 Nov 21];1–11. Available from: https://link.springer.com/article/10.1007/s00521-020-05626-8

15. Chinazzi M, Davis JT, Ajelli M, Gioannini C, Litvinova M, Merler S, et al. The effect of travel restrictions on the spread of the 2019 novel coronavirus (COVID-19) outbreak. Science (1979) [Internet]. 2020 Apr 24 [cited 2021 Dec 9];368(6489):395–400. Available from: https://www.science.org/doi/abs/10.1126/science.aba9757

16. Flaxman S, Mishra S, Gandy A, Unwin HJT, Mellan TA, Coupland H, et al. Estimating the effects of non-pharmaceutical interventions on COVID-19 in Europe. Nature [Internet]. 2020 Aug 13 [cited 2021 Dec 9];584(7820):257–61. Available from: https://pubmed.ncbi.nlm.nih.gov/32512579/

17. Tian H, Liu Y, Li Y, Wu CH, Chen B, Kraemer MUG, et al. An investigation of transmission control measures during the first 50 days of the COVID-19 epidemic in China. Science (1979) [Internet]. 2020 May 8 [cited 2021 Dec 9];368(6491):638–42. Available from: https://www.science.org/doi/abs/10.1126/science.abb6105

18. Lemaitre JC, Grantz KH, Kaminsky J, Meredith HR, Truelove SA, Lauer SA, et al. A scenario modeling pipeline for COVID-19 emergency planning. Scientific Reports 2021 11:1 [Internet]. 2021 Apr 6 [cited 2021 Jul 24];11(1):1–13. Available from: https://www.nature.com/articles/s41598-021-86811-0

19. Truelove S, Smith CP, Qin M, Mullany LC, Borchering RK, Lessler J, et al. Projected resurgence of COVID-19 in the United States in July—December 2021 resulting from the increased transmissibility of the Delta variant and faltering vaccination. medRxiv [Internet]. 2021 Sep 2 [cited 2022 Apr 9];16:2021.08.28.21262748. Available from: https://www.medrxiv.org/content/10.1101/2021.08.28.21262748v2

20. Borchering RK, Viboud C, Howerton E, Smith CP, Truelove S, Runge MC, et al. Modeling of Future COVID-19 Cases, Hospitalizations, and Deaths, by Vaccination Rates and Nonpharmaceutical Intervention Scenarios — United States, April–September 2021. Morbidity and Mortality Weekly Report [Internet]. 2021 [cited 2022 Apr 9];70(19):719. Available from: /pmc/articles/PMC8118153/

21. Chiu WA, Fischer R, Ndeffo-Mbah ML. State-level needs for social distancing and contact tracing to contain COVID-19 in the United States. Nature Human Behaviour 2020 4:10 [Internet]. 2020 Oct 6 [cited 2021 Dec 14];4(10):1080–90. Available from: https://www.nature.com/articles/s41562-020-00969-7

22. Luo J, Zhang Z, Fu Y, Rao F. Time series prediction of COVID-19 transmission in America using LSTM and XGBoost algorithms. Results in Physics. 2021 Aug 1;27:104462.

23. Duque D, Morton DP, Singh B, Du Z, Pasco R, Meyers LA. Timing social distancing to avert unmanageable COVID-19 hospital surges. Proc Natl Acad Sci U S A. 2020;117(33).

24. Shadabfar M, Mahsuli M, Sioofy Khoojine A, Hosseini VR. Time-variant reliability-based prediction of COVID-19 spread using extended SEIVR model and Monte Carlo sampling. Results in Physics. 2021;26.

25. Tkachenko A v., Maslov S, Elbanna A, Wong GN, Weiner ZJ, Goldenfeld N. Time-dependent heterogeneity leads to transient suppression of the COVID-19 epidemic, not herd immunity. Proc Natl Acad Sci U S A. 2021;118(17).

26. Shastri S, Singh K, Kumar S, Kour P, Mansotra V. Time series forecasting of Covid-19 using deep learning models: India-USA comparative case study. Chaos, Solitons & Fractals. 2020 Nov 1;140:110227.

27. Yan K, Yan H, Gupta R. The predicted trend of COVID-19 in the United States of America under the policy of “Opening Up America Again.” Infect Dis Model. 2021 Jan 1;6:766–81.

28. Guo X, Tong J, Chen P, Fan W. The suppression effect of emotional contagion in the COVID-19 pandemic: A multilayer hybrid modelling and simulation approach. PLoS ONE. 2021;16(7 July).

29. Lin YC, Chi WJ, Lin YT, Lai CY. The spatiotemporal estimation of the risk and the international transmission of COVID-19: a global perspective. Scientific Reports. 2020;10(1).

30. Bertozzi AL, Franco E, Mohler G, Short MB, Sledge D. The challenges of modeling and forecasting the spread of COVID-19. Proceedings of the National Academy of Sciences [Internet]. 2020 Jul 21 [cited 2021 Nov 2];117(29):16732–8. Available from: https://www.pnas.org/content/117/29/16732

31. Tang F, Feng Y, Chiheb H, Fan J. The Interplay of Demographic Variables and Social Distancing Scores in Deep Prediction of U.S. COVID-19 Cases. J Am Stat Assoc. 2021;116(534).

32. Alagoz O, Sethi AK, Patterson BW, Churpek M, Alhanaee G, Scaria E, et al. The impact of vaccination to control COVID-19 burden in the United States: A simulation modeling approach. PLoS ONE. 2021;16(7 July).

33. Hssayeni MD, Chala A, Dev R, Xu L, Shaw J, Furht B, et al. The forecast of COVID-19 spread risk at the county level. Journal of Big Data. 2021;8(1).

34. López L, Rodó X. The end of social confinement and COVID-19 re-emergence risk. Nature Human Behaviour. 2020;4(7).

35. Nande A, Sheen J, Walters EL, Klein B, Chinazzi M, Gheorghe AH, et al. The effect of eviction moratoria on the transmission of SARS-CoV-2. Nature Communications. 2021;12(1).

36. Bartsch SM, O’Shea KJ, Wedlock PT, Strych U, Ferguson MC, Bottazzi ME, et al. The Benefits of Vaccinating With the First Available COVID-19 Coronavirus Vaccine. American Journal of Preventive Medicine. 2021;60(5).

37. Wang Z, Zhang X, Teichert GH, Carrasco-Teja M, Garikipati K. System inference for the spatio-temporal evolution of infectious diseases: Michigan in the time of COVID-19. Computational Mechanics. 2020;66(5).

38. Chen S, Li Q, Gao S, Kang Y, Shi X. State-specific projection of COVID-19 infection in the United States and evaluation of three major control measures. Scientific Reports. 2020;10(1).

39. Unwin HJT, Mishra S, Bradley VC, Gandy A, Mellan TA, Coupland H, et al. State-level tracking of COVID-19 in the United States. Nature Communications. 2020;11(1).

40. Cuadros DF, Xiao Y, Mukandavire Z, Correa-Agudelo E, Hernández A, Kim H, et al. Spatiotemporal transmission dynamics of the COVID-19 pandemic and its impact on critical healthcare capacity. Health and Place. 2020;64.

41. Lawson AB, Kim J. Space-time covid-19 Bayesian SIR modeling in South Carolina. PLoS ONE. 2021;16(3 March).

42. Yasir KA, Liu WM. Social distancing mediated generalized model to predict epidemic spread of COVID-19. Nonlinear Dynamics. 2021;106(2).

43. Browning R, Sulem D, Mengersen K, Rivoirard V, Rousseau J. Simple discrete-time self-exciting models can describe complex dynamic processes: A case study of COVID-19. PLoS ONE. 2021;16(4 April).

44. Yu X, Lu L, Shen J, Li J, Xiao W, Chen Y. RLIM: a recursive and latent infection model for the prediction of US COVID-19 infections and turning points. Nonlinear Dynamics. 2021;106(2).

45. Vaid S, McAdie A, Kremer R, Khanduja V, Bhandari M. Risk of a second wave of Covid-19 infections: using artificial intelligence to investigate stringency of physical distancing policies in North America. International Orthopaedics. 2020;44(8).

46. Buckman SR, Glick R, Lansing KJ, Petrosky-Nadeau N, Seitelman LM. Replicating and projecting the path of COVID-19 with a model-implied reproduction number. Infect Dis Model. 2020;5.

47. Tsay C, Lejarza F, Stadtherr MA, Baldea M. Modeling, state estimation, and optimal control for the US COVID-19 outbreak. Scientific Reports 2020 10:1 [Internet]. 2020 Jul 1 [cited 2021 Nov 2];10(1):1–12. Available from: https://www.nature.com/articles/s41598-020-67459-8

48. Yamamoto N, Jiang B, Wang H. Quantifying compliance with COVID-19 mitigation policies in the US: A mathematical modeling study. Infect Dis Model. 2021;6.

49. Ilin C, Annan-Phan S, Tai XH, Mehra S, Hsiang S, Blumenstock JE. Public mobility data enables COVID-19 forecasting and management at local and global scales. Scientific Reports. 2021;11(1).

50. Shen M, Zu J, Fairley CK, Pagán JA, An L, Du Z, et al. Projected COVID-19 epidemic in the United States in the context of the effectiveness of a potential vaccine and implications for social distancing and face mask use. Vaccine. 2021;39(16).

51. Zhang X, Ma R, Wang L. Predicting turning point, duration and attack rate of COVID-19 outbreaks in major Western countries. Chaos, Solitons & Fractals. 2020 Jun 1;135:109829.

52. Watson GL, Xiong D, Zhang L, Zoller JA, Shamshoian J, Sundin P, et al. Pandemic velocity: Forecasting COVID-19 in the US with a machine learning & Bayesian time series compartmental model. PLOS Computational Biology [Internet]. 2021 Mar 1 [cited 2021 Nov 2];17(3):e1008837. Available from: https://journals.plos.org/ploscompbiol/article?id=10.1371/journal.pcbi.1008837

53. Renardy M, Eisenberg M, Kirschner D. Predicting the second wave of COVID-19 in Washtenaw County, MI. Journal of Theoretical Biology. 2020;507.

54. Hierro LÁ, Garzón AJ, Atienza-Montero P, Márquez JL. Predicting mortality for Covid-19 in the US using the delayed elasticity method. Scientific Reports. 2020;10(1).

55. Geng X, Gerges F, Katul GG, Bou-Zeid E, Nassif H, Boufadel MC. Population agglomeration is a harbinger of the spatial complexity of COVID-19. Chemical Engineering Journal. 2021;420.

56. Melin P, Sánchez D, Monica JC, Castillo O. Optimization using the firefly algorithm of ensemble neural networks with type-2 fuzzy integration for COVID-19 time series prediction. Soft Computing. 2021;

57. Cao Y, Francis R. On forecasting the community-level COVID-19 cases from the concentration of SARS-CoV-2 in wastewater. Science of the Total Environment. 2021;786.

58. Efimov D, Ushirobira R. On an interval prediction of COVID-19 development based on a SEIR epidemic model. Annual Reviews in Control. 2021;51.

59. Majid F, Gray M, Deshpande AM, Ramakrishnan S, Kumar M, Ehrlich S. Non-Pharmaceutical Interventions as Controls to mitigate the spread of epidemics: An analysis using a spatiotemporal PDE model and COVID–19 data. ISA Transactions. 2021;

60. Ekinci A. Modelling and forecasting of growth rate of new COVID-19 cases in top nine affected countries: Considering conditional variance and asymmetric effect. Chaos, Solitons and Fractals. 2021;151.

61. Reiner RC, Barber RM, Collins JK, Zheng P, Adolph C, Albright J, et al. Modeling COVID-19 scenarios for the United States. Nature Medicine 2020 27:1 [Internet]. 2020 Oct 23 [cited 2021 Nov 2];27(1):94–105. Available from: https://www.nature.com/articles/s41591-020-1132-9

62. Yang C, Wang J. Modeling the transmission of COVID-19 in the US – A case study. Infect Dis Model. 2021 Jan 1;6:195–211.

63. Wong GN, Weiner ZJ, Tkachenko A v., Elbanna A, Maslov S, Goldenfeld N. Modeling COVID-19 Dynamics in Illinois under Nonpharmaceutical Interventions. Physical Review X. 2020;10(4).

64. Singhal A, Singh P, Lall B, Joshi SD. Modeling and prediction of COVID-19 pandemic using Gaussian mixture model. Chaos, Solitons and Fractals. 2020;138.

65. Ala’raj M, Majdalawieh M, Nizamuddin N. Modeling and forecasting of COVID-19 using a hybrid dynamic model based on SEIRD with ARIMA corrections. Infect Dis Model. 2021;6.

66. Song J, Xie H, Gao B, Zhong Y, Gu C, Choi KS. Maximum likelihood-based extended Kalman filter for COVID-19 prediction. Chaos, Solitons and Fractals. 2021;146.

67. Sornette D, Mearns E, Schatz M, Wu K, Darcet D. Interpreting, analysing and modelling COVID-19 mortality data. Nonlinear Dynamics. 2020;101(3).

68. Liu M, Thomadsen R, Yao S. Forecasting the spread of COVID-19 under different reopening strategies. Scientific Reports 2020 10:1 [Internet]. 2020 Nov 23 [cited 2021 Nov 2];10(1):1–8. Available from: https://www.nature.com/articles/s41598-020-77292-8

69. Chalkiadakis I, Yan H, Peters GW, Shevchenko P v. Infection rate models for COVID-19: Model risk and public health news sentiment exposure adjustments. PLoS ONE. 2021;16(6 June).

70. Singh PK, Chouhan A, Bhatt RK, Kiran R, Ahmar AS. Implementation of the SutteARIMA method to predict short-term cases of stock market and COVID-19 pandemic in USA. Quality & Quantity. 2021;

71. España G, Cavany S, Oidtman R, Barbera C, Costello A, Lerch A, et al. Impacts of K-12 school reopening on the COVID-19 epidemic in Indiana, USA. Epidemics. 2021;37.

72. Cot C, Cacciapaglia G, Islind AS, Óskarsdóttir M, Sannino F. Impact of US vaccination strategy on COVID-19 wave dynamics. Scientific Reports. 2021;11(1).

73. Brugnago EL, da Silva RM, Manchein C, Beims MW. How relevant is the decision of containment measures against COVID-19 applied ahead of time? Chaos, Solitons and Fractals. 2020;140.

74. Khalilpourazari S, Hashemi Doulabi H, Özyüksel Çiftçioğlu A, Weber GW. Gradient-based grey wolf optimizer with Gaussian walk: Application in modelling and prediction of the COVID-19 pandemic. Expert Systems with Applications. 2021;177.

75. Basu S, Campbell RH. Going by the numbers: Learning and modeling COVID-19 disease dynamics. Chaos, Solitons and Fractals. 2020;138.

76. Singh P, Gupta A. Generalized SIR (GSIR) epidemic model: An improved framework for the predictive monitoring of COVID-19 pandemic. ISA Transactions. 2021;

77. Das RC. Forecasting incidences of COVID-19 using Box-Jenkins method for the period July 12-Septembert 11, 2020: A study on highly affected countries. Chaos, Solitons & Fractals. 2020 Nov 1;140:110248.

78. Feroze N. Forecasting the patterns of COVID-19 and causal impacts of lockdown in top five affected countries using Bayesian Structural Time Series Models. Chaos, Solitons & Fractals. 2020 Nov 1;140:110196.

79. Prasanth S, Singh U, Kumar A, Tikkiwal VA, Chong PHJ. Forecasting spread of COVID-19 using google trends: A hybrid GWO-deep learning approach. Chaos, Solitons & Fractals. 2021 Jan 1;142:110336.

80. Şahin U, Şahin T. Forecasting the cumulative number of confirmed cases of COVID-19 in Italy, UK and USA using fractional nonlinear grey Bernoulli model. Chaos, Solitons and Fractals. 2020;138.

81. Kalantari M. Forecasting COVID-19 pandemic using optimal singular spectrum analysis. Chaos, Solitons and Fractals. 2021;142.

82. Gecili E, Ziady A, Szczesniak RD. Forecasting COVID-19 confirmed cases, deaths and recoveries: Revisiting established time series modeling through novel applications for the USA and Italy. PLoS ONE. 2021;16(1 January).

83. da Silva RG, Ribeiro MHDM, Mariani VC, Coelho L dos S. Forecasting Brazilian and American COVID-19 cases based on artificial intelligence coupled with climatic exogenous variables. Chaos, Solitons and Fractals. 2020;139.

84. Nikolopoulos K, Punia S, Schäfers A, Tsinopoulos C, Vasilakis C. Forecasting and planning during a pandemic: COVID-19 growth rates, supply chain disruptions, and governmental decisions. European Journal of Operational Research. 2021;290(1).

85. Arias Velásquez RM, Mejía Lara JV. Forecast and evaluation of COVID-19 spreading in USA with reduced-space Gaussian process regression. Chaos, Solitons and Fractals. 2020;136.

86. Xu C, Yu Y, Chen YQ, Lu Z. Forecast analysis of the epidemics trend of COVID-19 in the USA by a generalized fractional-order SEIR model. Nonlinear Dynamics. 2020;101(3).

87. Salgotra R, Gandomi M, Gandomi AH. Evolutionary modelling of the COVID-19 pandemic in fifteen most affected countries. Chaos, Solitons & Fractals. 2020 Nov 1;140:110118.

88. Guo Y, Yu H, Zhang G, Ma DT. Exploring the impacts of travel-implied policy factors on COVID-19 spread within communities based on multi-source data interpretations. Health and Place. 2021;69.

89. Sharma RR, Kumar M, Maheshwari S, Ray KP. EVDHM-ARIMA-Based Time Series Forecasting Model and Its Application for COVID-19 Cases. IEEE Transactions on Instrumentation and Measurement. 2021;70.

90. Moghadas SM, Vilches TN, Zhang K, Nourbakhsh S, Sah P, Fitzpatrick MC, et al. Evaluation of COVID-19 vaccination strategies with a delayed second dose. PLoS Biology. 2021;19(4).

91. Kuo CP, Fu JS. Evaluating the impact of mobility on COVID-19 pandemic with machine learning hybrid predictions. Science of the Total Environment. 2021;758.

92. Li Q, Bedi T, Lehmann CU, Xiao G, Xie Y. Evaluating short-term forecasting of COVID-19 cases among different epidemiological models under a Bayesian framework. Gigascience. 2021;10(2).

93. Mahajan A, Solanki R, Sivadas N. Estimation of undetected symptomatic and asymptomatic cases of COVID-19 infection and prediction of its spread in the USA. Journal of Medical Virology. 2021;93(5).

94. Lee SY, Lei B, Mallick B. Estimation of COVID-19 spread curves integrating global data and borrowing information. PLoS ONE. 2020;15(7 July).

95. Levin MW, Shang M, Stern R. Effects of short-term travel on COVID-19 spread: A novel SEIR model and case study in Minnesota. PLoS ONE. 2021;16(1 January).

96. Wang X, Du Z, Johnson KE, Pasco RF, Fox SJ, Lachmann M, et al. Effects of COVID-19 vaccination timing and risk prioritization on mortality rates, United States. Emerging Infectious Diseases. 2021;27(7).

97. Tam KM, Walker N, Moreno J. Effect of mitigation measures on the spreading of COVID-19 in hard-hit states in the U.S. PLoS ONE. 2020;15(11 November).

98. Zeng X, Ghanem R. Dynamics identification and forecasting of COVID-19 by switching Kalman filters. Computational Mechanics. 2020;66(5).

99. Kirpich A, Koniukhovskii V, Shvartc V, Skums P, Weppelmann TA, Imyanitov E, et al. Development of an interactive, agent-based local stochastic model of COVID-19 transmission and evaluation of mitigation strategies illustrated for the state of Massachusetts, USA. PLoS ONE. 2021;16(2 Febuary).

100. Zeroual A, Harrou F, Dairi A, Sun Y. Deep learning methods for forecasting COVID-19 time-Series data: A Comparative study. Chaos, Solitons & Fractals. 2020 Nov 1;140:110121.

101. Shirin A, Lin YT, Sorrentino F. Data-driven optimized control of the COVID-19 epidemics. Scientific Reports. 2021;11(1).

102. Bhardwaj R, Bangia A. Data driven estimation of novel COVID-19 transmission risks through hybrid soft-computing techniques. Chaos, Solitons & Fractals. 2020 Nov 1;140:110152.

103. Ballı S. Data analysis of Covid-19 pandemic and short-term cumulative case forecasting using machine learning time series methods. Chaos, Solitons & Fractals. 2021 Jan 1;142:110512.

104. Chen S, Chen Q, Yang J, Lin L, Li L, Jiao L, et al. Curbing the COVID-19 pandemic with facility-based isolation of mild cases: a mathematical modeling study. J Travel Med. 2021;28(2).

105. Zhao H, Merchant NN, McNulty A, Radcliff TA, Cote MJ, Fischer RSB, et al. COVID-19: Short term prediction model using daily incidence data. PLoS ONE. 2021;16(4 April).

106. Pacheco-Barrios K, Cardenas-Rojas A, Giannoni-Luza S, Fregni F. COVID-19 pandemic and Farr’s law: A global comparison and prediction of outbreak acceleration and deceleration rates. PLoS ONE. 2020;15(9 September).

107. Gel ES, Jehn M, Lant T, Muldoon AR, Nelson T, Ross HM. COVID-19 healthcare demand projections: Arizona. PLoS ONE. 2020;15(12 December).

108. Bhouri MA, Costabal FS, Wang H, Linka K, Peirlinck M, Kuhl E, et al. COVID-19 dynamics across the US: A deep learning study of human mobility and social behavior. Computer Methods in Applied Mechanics and Engineering. 2021;382.

109. Kerr CC, Stuart RM, Mistry D, Abeysuriya RG, Rosenfeld K, Hart GR, et al. Covasim: An agent-based model of COVID-19 dynamics and interventions. PLoS Computational Biology. 2021;17(7).

110. Er S, Yang S, Zhao T. COUnty aggRegation mixup AuGmEntation (COURAGE) COVID-19 prediction. Scientific Reports. 2021;11(1).

111. Chan S, Chu J, Zhang Y, Nadarajah S. Count regression models for COVID-19. Physica A: Statistical Mechanics and its Applications. 2021;563.

112. Chen X, Zhang A, Wang H, Gallaher A, Zhu X. Compliance and containment in social distancing: mathematical modeling of COVID-19 across townships. International Journal of Geographical Information Science. 2021;35(3).

113. Dairi A, Harrou F, Zeroual A, Hittawe MM, Sun Y. Comparative study of machine learning methods for COVID-19 transmission forecasting. Vol. 118, Journal of Biomedical Informatics. 2021.

114. Zebrowski A, Rundle A, Pei S, Yaman T, Yang W, Carr BG, et al. A Spatiotemporal Tool to Project Hospital Critical Care Capacity and Mortality From COVID-19 in US Counties. https://doi.org/102105/AJPH2021306220 [Internet]. 2021 May 5 [cited 2021 Nov 2];111(6):1113–22. Available from: https://ajph.aphapublications.org/doi/abs/10.2105/AJPH.2021.306220

115. Yan H, Zhu Y, Gu J, Huang Y, Sun H, Zhang X, et al. Better strategies for containing COVID-19 pandemic: A study of 25 countries via a vSIADR model. Proceedings of the Royal Society A: Mathematical, Physical and Engineering Sciences. 2021;477(2248).

116. Patel MD, Rosenstrom E, Ivy JS, Mayorga ME, Keskinocak P, Boyce RM, et al. Association of Simulated COVID-19 Vaccination and Nonpharmaceutical Interventions with Infections, Hospitalizations, and Mortality. JAMA Network Open. 2021;

117. Yu D, Zhu G, Wang X, Zhang C, Soltanalizadeh B, Wang X, et al. Assessing effects of reopening policies on COVID-19 pandemic in Texas with a data-driven transmission model. Infect Dis Model. 2021;6.

118. Zhang Y, Yu X, Sun HG, Tick GR, Wei W, Jin B. Applicability of time fractional derivative models for simulating the dynamics and mitigation scenarios of COVID-19. Chaos, Solitons and Fractals. 2020;138.

119. Naz R, Al-Raeei M. Analysis of transmission dynamics of COVID-19 via closed-form solutions of a susceptible-infectious-quarantined-diseased model with a quarantine-adjusted incidence function. Mathematical Methods in the Applied Sciences. 2021;44(14).

120. Nadler P, Wang S, Arcucci R, Yang X, Guo Y. An epidemiological modelling approach for COVID-19 via data assimilation. European Journal of Epidemiology. 2020;35(8).

121. Shamil MS, Farheen F, Ibtehaz N, Khan IM, Rahman MS. An Agent-Based Modeling of COVID-19: Validation, Analysis, and Recommendations. Cognitive Computation. 2021;

122. Upadhyay RK, Chatterjee S, Saha S, Azad RK. Age-group-targeted testing for COVID-19 as a new prevention strategy. Nonlinear Dynamics. 2020;101(3).

123. Ramazi P, Haratian A, Meghdadi M, Mari Oriyad A, Lewis MA, Maleki Z, et al. Accurate long-range forecasting of COVID-19 mortality in the USA. Scientific Reports. 2021;11(1).

124. Muñoz-Fernández GA, Seoane JM, Seoane-Sepúlveda JB. A SIR-type model describing the successive waves of COVID-19. Chaos, Solitons and Fractals. 2021;144.

125. Cooper I, Mondal A, Antonopoulos CG. A SIR model assumption for the spread of COVID-19 in different communities. Chaos, Solitons and Fractals. 2020;139.

126. Koutsellis T, Nikas A. A predictive model and country risk assessment for COVID-19: An application of the Limited Failure Population concept. Chaos, Solitons and Fractals. 2020;140.

127. Ren J, Yan Y, Zhao H, Ma P, Zabalza J, Hussain Z, et al. A Novel Intelligent Computational Approach to Model Epidemiological Trends and Assess the Impact of Non-Pharmacological Interventions for COVID-19. IEEE Journal of Biomedical and Health Informatics. 2020;24(12).

128. Ramezani SB, Amirlatifi A, Rahimi S. A novel compartmental model to capture the nonlinear trend of COVID-19. Computers in Biology and Medicine. 2021;134.

129. Mokhtari A, Mineo C, Kriseman J, Kremer P, Neal L, Larson J. A multi-method approach to modeling COVID-19 disease dynamics in the United States. Scientific Reports. 2021;11(1).

130. Usherwood T, LaJoie Z, Srivastava V. A model and predictions for COVID-19 considering population behavior and vaccination. Scientific Reports. 2021;11(1).

131. Rastgoftar H, Atkins E. A Mass-Conservation Model for Stability Analysis and Finite-Time Estimation of Spread of COVID-19. IEEE Transactions on Computational Social Systems. 2021;8(4).

132. Lu Z, Yu Y, Chen YQ, Ren G, Xu C, Wang S, et al. A fractional-order SEIHDR model for COVID-19 with inter-city networked coupling effects. Vol. 101, Nonlinear Dynamics. 2020.

133. Ertem Z, Araz OM, Cruz-Aponte M. A decision analytic approach for social distancing policies during early stages of COVID-19 pandemic. Decision Support Systems. 2021;

134. Paiva HM, Afonso RJM, de Oliveira IL, Garcia GF. A data-driven model to describe and forecast the dynamics of COVID-19 transmission. PLoS ONE. 2020;15(7 July).

135. Hasan A, Nasution Y. A compartmental epidemic model incorporating probable cases to model COVID-19 outbreak in regions with limited testing capacity. ISA Transactions. 2021.

136. Brethouwer JT, van de Rijt A, Lindelauf R, Fokkink R. “Stay nearby or get checked”: A Covid-19 control strategy. Infect Dis Model. 2021;6.

137. Pei S, Kandula S, Shaman J. Differential effects of intervention timing on COVID-19 spread in the United States. Science Advances. 2020 Dec 4;6(49).

138. Khan ZS, van Bussel F, Hussain F. A predictive model for Covid-19 spread applied to eight US states. 2020 Jun 10 [cited 2021 Nov 2]; Available from: https://arxiv.org/abs/2006.05955v4

139. Galasso J, Cao DM, Hochberg R. A random forest model for forecasting regional COVID-19 cases utilizing reproduction number estimates and demographic data. medRxiv [Internet]. 2021 Sep 14 [cited 2021 Nov 2];2021.05.23.21257689. Available from: https://www.medrxiv.org/content/10.1101/2021.05.23.21257689v2

140. Zhang-James Y, Hess J, Salekin A, Wang D, Chen S, Winkelstein P, et al. A seq2seq model to forecast the COVID-19 cases, deaths and reproductive R numbers in US counties. medRxiv [Internet]. 2021 Apr 20 [cited 2021 Nov 2];2021.04.14.21255507. Available from: https://www.medrxiv.org/content/10.1101/2021.04.14.21255507v1

141. Rowland MA, Swannack TM, Mayo ML, Parno M, Farthing M, Dettwiller I, et al. COVID-19 infection data encode a dynamic reproduction number in response to policy decisions with secondary wave implications. Scientific Reports 2021 11:1 [Internet]. 2021 May 25 [cited 2021 Nov 2];11(1):1–7. Available from: https://www.nature.com/articles/s41598-021-90227-1

142. Rodriguez A, Tabassum A, Cui J, Xie J, Ho J, Agarwal P, et al. DeepCOVID: An Operational Deep Learning-driven Framework for Explainable Real-time COVID-19 Forecasting. medRxiv [Internet]. 2020 Sep 29 [cited 2021 Nov 2];2020.09.28.20203109. Available from: https://www.medrxiv.org/content/10.1101/2020.09.28.20203109v1

143. Biegel HR, Lega J. EpiCovDA: a mechanistic COVID-19 forecasting model with data assimilation. ArXiv. 2021.

144. Zou D, Wang L, Xu P, Chen J, Zhang W, Gu Q. Epidemic Model Guided Machine Learning for COVID-19 Forecasts in the United States. medRxiv [Internet]. 2020 May 25 [cited 2021 Nov 2];2020.05.24.20111989. Available from: https://www.medrxiv.org/content/10.1101/2020.05.24.20111989v1

145. Srivastava A, Xu T, Prasanna VK. Fast and Accurate Forecasting of COVID-19 Deaths Using the SIkJα Model. 2020 Jul 10 [cited 2021 Nov 2]; Available from: https://arxiv.org/abs/2007.05180v2

146. team IC 19 health service utilization forecasting, Murray CJ. Forecasting COVID-19 impact on hospital bed-days, ICU-days, ventilator-days and deaths by US state in the next 4 months. medRxiv [Internet]. 2020 Mar 30 [cited 2021 Nov 2];2020.03.27.20043752. Available from: https://www.medrxiv.org/content/10.1101/2020.03.27.20043752v1

147. Pei S, Shaman J. Initial Simulation of SARS-CoV2 Spread and Intervention Effects in the Continental US. medRxiv [Internet]. 2020 Mar 27 [cited 2021 Nov 2];2020.03.21.20040303. Available from: https://www.medrxiv.org/content/10.1101/2020.03.21.20040303v2

148. Arik SO, Li CL, Yoon J, Sinha R, Epshteyn A, Le LT, et al. Interpretable Sequence Learning for COVID-19 Forecasting. Advances in Neural Information Processing Systems [Internet]. 2020 Aug 3 [cited 2021 Nov 2];2020-December. Available from: https://arxiv.org/abs/2008.00646v2

149. Neural Relational Autoregression for High-Resolution COVID-19 Forecasting [Internet]. [cited 2021 Nov 2]. Available from: https://ai.facebook.com/research/publications/neural-relational-autoregression-for-high-resolution-covid-19-forecasting/

150. Gibson GC, Reich NG, Sheldon D. Real-time Mechanistic Bayesian Forecasts of COVID-19 Mortality. medRxiv [Internet]. 2020 [cited 2021 Nov 2]; Available from: /pmc/articles/PMC7781348/

151. Gao J, Sharma R, Qian C, Glass LM, Spaeder J, Romberg J, et al. STAN: spatio-temporal attention network for pandemic prediction using real-world evidence. Journal of the American Medical Informatics Association [Internet]. 2021 Mar 18 [cited 2021 Nov 2];28(4):733–43. Available from: https://academic.oup.com/jamia/article/28/4/733/6118380

152. Baxter A, Oruc BE, Keskinocak P, Asplund J, Serban N. Evaluating Scenarios for School Reopening under COVID19. medRxiv [Internet]. 2020 Jul 24 [cited 2021 Dec 14];2020.07.22.20160036. Available from: https://www.medrxiv.org/content/10.1101/2020.07.22.20160036v1

153. Wang L, Wang G, Gao L, Li X, Yu S, Kim M, et al. Spatiotemporal Dynamics, Nowcasting and Forecasting of COVID-19 in the United States. 2020 Apr 29 [cited 2021 Dec 14]; Available from: https://arxiv.org/abs/2004.14103v4

154. Wilson DJ, thank Regis Barnichon I, Leduc S, Mertens K, Moretti E, Roth Tran B. Weather, Mobility, and COVID-19: A Panel Local Projections Estimator for Understanding and Forecasting Infectious Disease Spread. Federal Reserve Bank of San Francisco [Internet]. 2020 [cited 2022 Apr 1]; Available from: https://doi.org/10.24148/wp2020-23

155. Shi Y, Ban X. Capping Mobility to Control COVID-19: A Collision-based Infectious Disease Transmission Model. medRxiv [Internet]. 2020 Jul 28 [cited 2021 Nov 2];2020.07.25.20162016. Available from: https://www.medrxiv.org/content/10.1101/2020.07.25.20162016v1

156. Wu D, Gao L, Xiong X, Chinazzi M, Vespignani A, Ma YA, et al. DeepGLEAM: A hybrid mechanistic and deep learning model for COVID-19 forecasting. 2021 Feb 12 [cited 2022 Apr 15]; Available from: https://arxiv.org/abs/2102.06684v3

157. Pollett S, Johansson MA, Reich NG, Brett-Major D, del Valle SY, Venkatramanan S, et al. Recommended reporting items for epidemic forecasting and prediction research: The EPIFORGE 2020 guidelines. PLOS Medicine [Internet]. 2021 Oct 1 [cited 2021 Dec 6];18(10):e1003793. Available from: https://journals.plos.org/plosmedicine/article?id=10.1371/journal.pmed.1003793

158. COVID-19 Community Mobility Reports [Internet]. [cited 2021 Dec 14]. Available from: https://www.google.com/covid19/mobility/

159. Covid-19 Social Distancing Scoreboard — Unacast [Internet]. [cited 2021 Dec 14]. Available from: https://www.unacast.com/covid19/social-distancing-scoreboard

160. SafeGraph | Academics [Internet]. [cited 2021 Dec 14]. Available from: https://www.safegraph.com/academics

161. COVID-19 - Mobility Trends Reports - Apple [Internet]. [cited 2021 Dec 14]. Available from: https://covid19.apple.com/mobility

162. Facebook Data For Good Movement Range Maps [Internet]. [cited 2021 Dec 14]. Available from: https://dataforgood.facebook.com/dfg/tools/movement-range-maps

163. Coronavirus Search Trends - Google Trends [Internet]. [cited 2021 Dec 14]. Available from: https://trends.google.com/trends/story/GB_cu_JSW_pHABAADqAM_en

164. COVID-19 [Internet]. [cited 2021 Dec 14]. Available from: https://covid19.healthdata.org/global?view=mask-use&tab=trend

165. Facebook Data For Good COVID 19 Symptom Survey [Internet]. [cited 2021 Dec 14]. Available from: https://dataforgood.facebook.com/dfg/tools/covid-19-trends-and-impact-survey#methodology

166. covid-19-data/mask-use at master · nytimes/covid-19-data · GitHub [Internet]. [cited 2021 Dec 14]. Available from: https://github.com/nytimes/covid-19-data/tree/master/mask-use

167. Hale T, Angrist N, Goldszmidt R, Kira B, Petherick A, Phillips T, et al. A global panel database of pandemic policies (Oxford COVID-19 Government Response Tracker). Nature Human Behaviour 2021 5:4 [Internet]. 2021 Mar 8 [cited 2021 Dec 9];5(4):529–38. Available from: https://www.nature.com/articles/s41562-021-01079-8

168. Adolph C, Amano K, Bang-Jensen B, Fullman N, Wilkerson J. Pandemic Politics: Timing State-Level Social Distancing Responses to COVID-19. Journal of Health Politics, Policy and Law. 2021 Apr 1;46(2):211–33.

169. Lutz CS, Huynh MP, Schroeder M, Anyatonwu S, Dahlgren FS, Danyluk G, et al. Applying infectious disease forecasting to public health: A path forward using influenza forecasting examples. BMC Public Health [Internet]. 2019 Dec 10 [cited 2021 Dec 8];19(1):1–12. Available from: https://bmcpublichealth.biomedcentral.com/articles/10.1186/s12889-019-7966-8

170. Home - COVID 19 forecast hub [Internet]. [cited 2021 Dec 7]. Available from: https://covid19forecasthub.org/eval-reports/?state=US&week=2021-09-08

171. Johansson MA, Reich NG, Meyers LA, Lipsitch M. Preprints: An underutilized mechanism to accelerate outbreak science. PLOS Medicine [Internet]. 2018 Apr 1 [cited 2021 Dec 14];15(4):e1002549. Available from: https://journals.plos.org/plosmedicine/article?id=10.1371/journal.pmed.1002549

172. 2019 Novel Coronavirus Research Compendium (NCRC) [Internet]. [cited 2021 Dec 8]. Available from: https://ncrc.jhsph.edu/

173. Gardner L, Ratcliff J, Dong E, Katz A. A need for open public data standards and sharing in light of COVID-19. The Lancet Infectious Diseases [Internet]. 2021 Apr 1 [cited 2021 Dec 9];21(4):e80. Available from: http://www.thelancet.com/article/S1473309920306356/fulltext

174. Reinhart A, Brooks L, Jahja M, Rumack A, Tang J, Agrawal S, et al. An Open Repository of Real-Time COVID-19 Indicators. medRxiv [Internet]. 2021 Nov 11 [cited 2021 Dec 9];2021.07.12.21259660. Available from: https://www.medrxiv.org/content/10.1101/2021.07.12.21259660v2

175. Badr HS, D. H, Marshall M, Dong E, Squire MM, Gardner LM. Association between mobility patterns and COVID-19 transmission in the USA: a mathematical modelling study. The Lancet Infectious Diseases. 2020 Nov 1;20(11):1247–54.

176. Pollett S, Johansson MA, Reich NG, Brett-Major D, del Valle SY, Venkatramanan S, et al. Recommended reporting items for epidemic forecasting and prediction research: The EPIFORGE 2020 guidelines. PLOS Medicine [Internet]. 2021 Oct 1 [cited 2021 Dec 8];18(10):e1003793. Available from: https://journals.plos.org/plosmedicine/article?id=10.1371/journal.pmed.1003793

177. Meredith HR, Arehart E, Grantz KH, Beams A, Sheets T, Nelson R, et al. Coordinated Strategy for a Model-Based Decision Support Tool for Coronavirus Disease, Utah, USA. Emerging Infectious Diseases [Internet]. 2021 May 1 [cited 2021 Dec 9];27(5):1259. Available from: /pmc/articles/PMC8084489/

